# A Software Platform for Collaborative Infectious Disease Modeling

**DOI:** 10.1101/2025.10.03.25337284

**Authors:** Consortium of Infectious Disease Modeling Hubs, Melissa Kerr, Rebecca Borchering, Alvaro Castro Rivadeneira, Lucie Contamin, Sebastian Funk, Harry Hochheiser, Emily Howerton, Anna Krystalli, Li Shandross, Nicholas G Reich

## Abstract

To better respond to threats, decision-makers are increasingly interested in predictions they can understand and trust. Collaborative modeling can help increase the relevance, transparency, and robustness of predictions. This approach can be facilitated with hubs, or centralized data repositories to collect, analyze, and communicate model output. This paper introduces the hubverse, a suite of standards and software tools to streamline the creation and operation of collaborative modeling hubs. Hubverse file structure and model output standards enable the use of common tools to validate, aggregate, visualize, evaluate, and communicate model output. Currently, the hubverse is used by nearly two dozen collaborative and local modeling hubs around the globe to support infectious disease modeling efforts, including hubs hosted and/or used by the United States Centers for Disease Control and Prevention, European Centre for Disease Prevention and Control, Australia-Aotearoa Consortium for Epidemic Forecasting and Analytics, and California Department of Public Health.

---

Reliable predictions can help decision-makers respond to emerging public health threats. Predictive modeling has played a prominent role in initiating public health response to Ebola^1^ and informing COVID-19 restrictive measures^2^ and vaccination strategies^3^. As efforts to integrate predictive modeling into real-time decision-making increase^4,5^, effective communication between modelers and decision-makers is necessary to ensure the latter have model output that they can understand, trust, and use.

However, increased infectious disease modeling efforts have also resulted in a complex landscape for decision-makers, characterized by a wide range of predicted outcomes, varying public health relevance, and diverse evaluation metrics^6^. These factors complicate model comparison and make it difficult to select the best model to guide decisions^6^. To address these challenges, coordinated multi-model approaches have been adopted to improve decision-making, including long-term policy-focused consortia (e.g., Vaccine Impact Modelling Consortium^7^, TB Modelling and Analysis Consortium^8^, Malaria Consortium^9^) and related studies evaluating vaccine impact^10–12^, alongside real-time forecasting collaborations for influenza^13^, COVID-19^14^, dengue^15^, and Ebola^16^. Multi-model approaches facilitate model comparison, better acknowledge uncertainty, and identify patterns across differing assumptions and methods^17^.

By bringing together predictions from multiple, independently developed models, multi-model efforts lay the foundation for ensemble modeling. In aggregating predictions from multiple models, ensembles provide a more robust, comprehensive understanding of potential futures or scenarios and help mitigate the risks associated with model-specific biases or limitations^18^. When individual models offer diverse insight on potential future outcomes, ensemble approaches can better capture uncertainty and yield more accurate forecasts than any single model^19^.

This improved performance has been demonstrated across a range of outbreak settings, including influenza^20^, COVID-19^21,22^, Ebola^16^, and dengue^15^, even when using simple, untrained ensembling approaches such as equal-weight mean or median ensembles^21,23,24^. In some settings, accounting for past model performance and fully incorporating uncertainty can further enhance ensemble performance^23,25^.

Beyond predictive gains, ensemble models provide more reliable model output for decision-makers, which often makes them the primary hub product communicated^21,26,27^. For example, ensemble projections generated by the U.S. COVID-19 Scenario Modeling Hub helped support decisions to broaden the COVID-19 vaccination program to 5-11 year olds^28^, hasten the distribution of bivalent vaccines in response to circulating variants^3^, and administer boosters to a larger portion of the population^29^.

To facilitate ensemble building and improve the overall utility of modeling, collaborative modeling hubs can be established. We define collaborative modeling as a research consortium that responds to a scientific challenge through coordinated modeling efforts^18^. We define a hub as a centralized data repository to collect, analyze, and communicate model predictions, i.e., *model output*. Collaborative modeling hubs offer many benefits. First, by imposing model output standards, they make it easier to build ensemble models. Second, hubs may serve as a central communication point between modelers and stakeholders, helping to ensure that the outcomes being predicted and the model output generated meet decision-making needs^30^. Third, in eliciting model output submissions in response to a common challenge, hubs improve scientific rigor by encouraging model output transparency, data and methods sharing, and comparisons across models^18^. Furthermore, while often established as multi-institutional collaborations, it is important to note that modeling hubs can also be implemented within a single research group or laboratory to support structured model comparison, internal benchmarking, ensemble development, and methodological innovation.

One of the earliest collaborative modeling hubs was the FluSight influenza challenge, launched by the U.S. Centers for Disease Control and Prevention (U.S. CDC) in 2013 to forecast influenza trends 1-4 weeks into the future to improve risk assessment and preparedness. FluSight introduced quantitative forecasting standards and evaluation and cultivated contributions from academia and industry^26^. When the COVID-19 pandemic hit, experience from FluSight helped inform the U.S. COVID-19 Forecast Hub. Launched in April 2020 in response to the massive number of forecasts generated during the early pandemic, the U.S. COVID-19 Forecast Hub sought to standardize 1-4 week-ahead forecasts of incident cases, deaths, and hospitalizations so that model output could be compared, aggregated into ensemble models, and communicated to stakeholders^31^.

The hubverse^32^, the topic of the current paper, builds off the efforts of these, and other similar, infectious disease forecasting hubs by introducing standards and tools to streamline collaborative modeling efforts and make it easier to set up hubs and communicate model output. The hubverse introduces file structure and model output standards, which enable the use of tools to validate model submissions and aggregate, visualize, evaluate, and communicate model output.

## Results

### Summary of hubverse architecture

A hub is a repository that contains model output and metadata files that conform to specific standards. All data are stored in a version-controlled repository. Configuration files (JSON text files) in the hub configuration directory define how the hub is set up and the scientific challenge, or *modeling tasks*, to be addressed.

Hubs solicit model output submissions from modeling teams during specific time periods, or *rounds*, to address modeling tasks. For example, hubs may solicit model output once a week to predict weekly counts of hospital admissions due to influenza. Model output is submitted as individual files of tabular data to a hub’s model output directory, with one file representing one model and one round. Hubs may collect point predictions, which provide a single numerical estimate, or probabilistic predictions, which assign a likelihood to a range of outcomes. The latter are often preferred by decision-makers and modelers in diverse fields since they provide a better representation of risk and uncertainty^33–35^.

For some modeling tasks, predicted values of an outcome can be evaluated against actual observed values (e.g., forecasts of hospitalization trends can later be evaluated against actual observed hospitalizations). These actual observed values, referred to as *target data*, may be stored in a hub’s target data directory.

Hubverse data standards ensure that model output data can be seamlessly integrated to easily validate, aggregate, visualize, and evaluate predictions (**Figure 1**).

**Figure 1.**
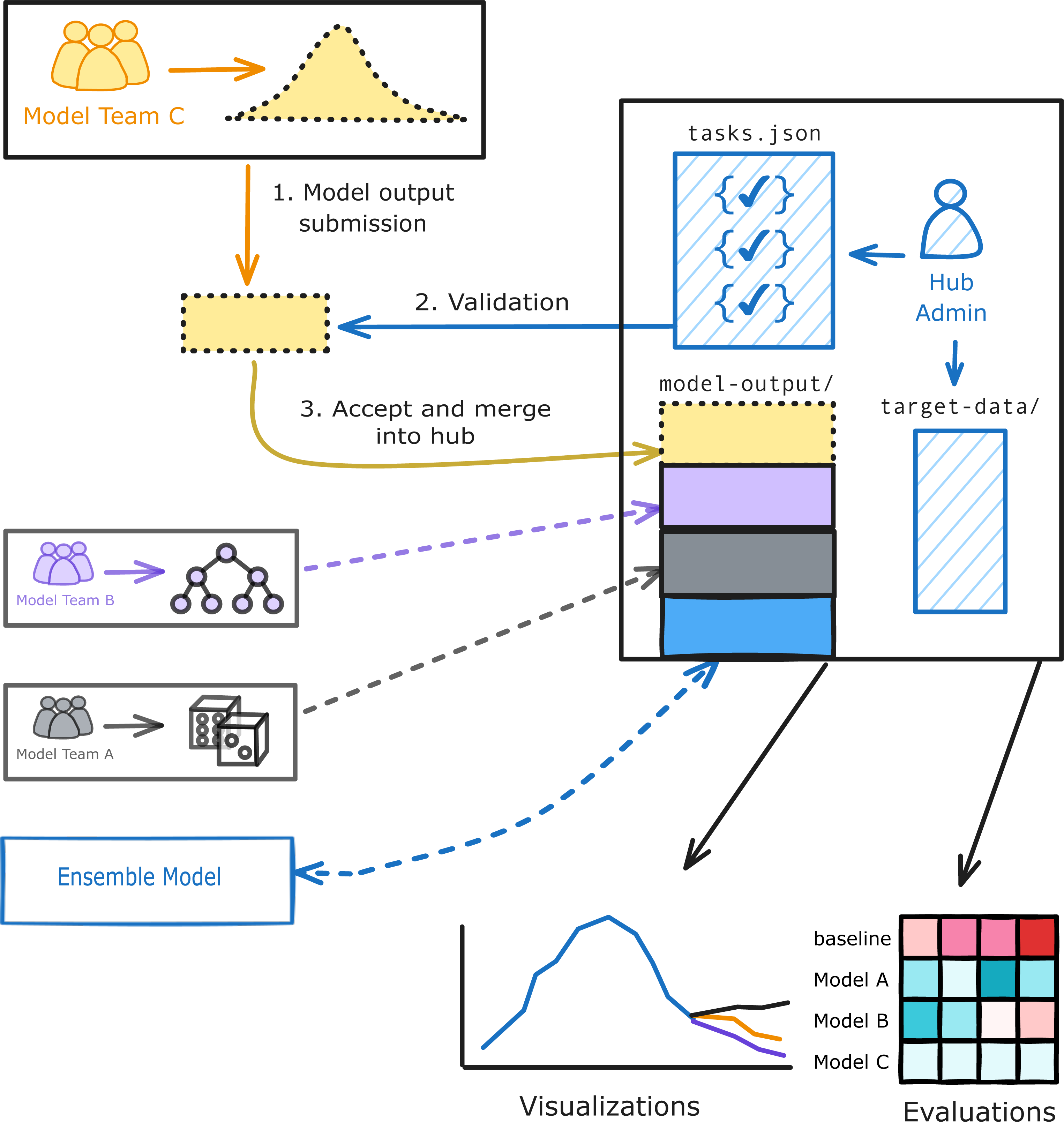
Overview of hub architecture Modeling tasks are defined in the tasks.json configuration file. Model output submissions are collected in the model-output directory of a hub after being validated against the tasks.json configuration file. The optional target-data directory contains actual observed values of an event.

### Examples from the hubverse community

Hubverse infrastructure was developed by the Consortium of Infectious Disease Modeling Hubs, a team of software developers and researchers from academia with experience building and managing hubs who came together to streamline efforts following the COVID-19 pandemic. Since its inception in 2022, hubverse infrastructure has been adopted by groups both outside and inside academia. Collaborative hubs using hubverse infrastructure have been established around the globe to collect predictions from diverse modeling teams for a range of infectious diseases, including respiratory illnesses and arboviruses (**Extended Data Table 1)**. Many of these hubs can be denoted as *community hubs* that have public repositories and are often open to participation from any modeling team.

Hubverse infrastructure is also used by individual modelers or teams to build *local hubs,* i.e., hubs set up on a laptop or cluster. Local hubs can support model development for general research purposes or future submission to a community modeling hub. For example, a modeler could run and store model output in the local hub, and then use hubverse validation, visualization, and evaluation tools to analyze different models. Running and comparing different model variants in a controlled experimental setting allows researchers to more formally measure and report uncertainties due to model choices and assumptions, thereby increasing model transparency and robustness.

### How different users interact with hubverse tools at different phases of a hub

The hubverse contributes to public health through the collaboration between users in different roles, of which we define five: the hub administrator, modeler, analyst, developer, and stakeholder (**Figure 2**). We note that these are not mutually exclusive and a single person might serve in multiple roles, or a single role might be filled by multiple people.

**Figure 2.**
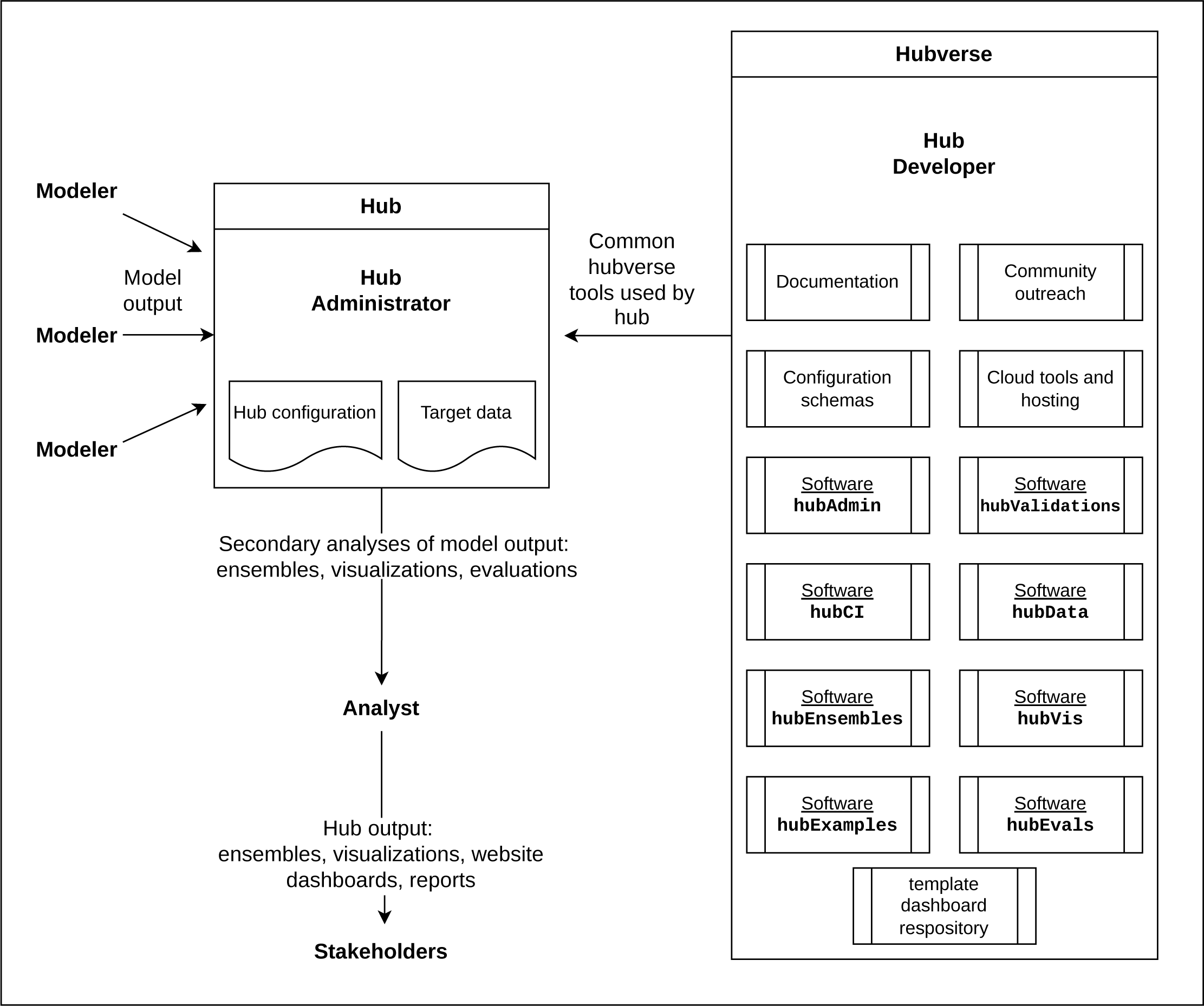
Schematic of the hubverse showing different user roles. The ecosystem of hubverse tools is shown in the box on the right. The ecosystem includes tools to set up and administer a hub, as well as tools to access, validate, aggregate, and visualize model output. A hub that follows the hubverse template is shown in the middle, with modelers contributing model output, hub administrators coordinating internal hub activities, and analysts processing model outputs into secondary products for stakeholders to consult.

Different user roles typically interact with the hub at different times. The lifecycle of a hub is organized around *rounds*, or specific time periods during which model output is solicited in response to a scientific challenge, or *modeling tasks*. The definition of a round may differ based on the hub. For example, hubs that accept daily submissions might consider each day a separate round, while other hubs may have a round every week, month, or season, with a submission period open for multiple days. Based on this concept of a round, we define four phases in the lifecycle of a hub: 1) hub configuration, 2) active round, 3) post-round analysis, and 4) stakeholder products. In the following subsections we describe the ways in which users interact with hubverse tools and products at different hub phases. **Table 1** lists the hubverse tools used by different users at each hub phase. **Supplementary Figure 1** provides a flowchart of the hub phases and the tools used in each phase.

**Table 1.** Structured overview of how different users interact with hubverse tools or products at each hub phase.

| Phase | Tool | User |  |  |
| --- | --- | --- | --- | --- |
| Hub set-up |  | Admin | Modeler | Analyst |
| Hosting/storing hub data | GitHub repositories, AWS buckets | X |  |  |
| Setting up a hub repository | Website tutorial, hubTemplate repository | X |  |  |
| Creating configuration files | Website tutorial, hubAdmin | X |  |  |
| Validating configuration files | hubAdmin | X |  |  |
| Defining modeling tasks conceptually | Website offers terminology and guidance, hubs need to customize | X |  |  |
| Setting up continuous integration | hubCI, GitHub Action templates | X |  |  |
| **Active round** |  |  |  |  |
| Storing target data snapshots | Hub repository (optional) | X |  |  |
| Submitting model output | Hub repository |  | X |  |
| Submitting model metadata | Hub repository |  | X |  |
| Validating model submissions | hubCI, hubValidations | X | X |  |
| **Post-round analysis** |  |  |  |  |
| Connecting to model output data | hubData |  |  | X |
| Building an ensemble | hubEnsembles |  |  | X |
| Visualizing data | hubVis, hubExamples |  | X | X |
| Scoring model output data | hubEvals |  | X | X |
| Building reports | hubVis, hubEvals | X |  | X |
| Building a website/dashboard | hubVis, hubEvals, hub template dashboard repository | X |  | X |

The hubverse landing page (https://hubverse.io) and documentation site (https://docs.hubverse.io/) provide access to hubverse data standards, tools, and documentation. Most packages listed in the sections below have vignettes accessible through links provided in the Data Availability section.

### Hub administrators

Hub administrators are involved in hub set-up and the day-to-day management of a hub. Hub administrators decide where to host and store hub data. The hubverse currently supports primary data storage on GitHub with an optional backup to Amazon Web Services Simple Storage Service (AWS S3). To set up a hub repository on GitHub, the hubverse website provides tutorials and template repositories. Template repositories include the three required configuration files (admin.json, tasks.json, and model-metadata-schema.json) that administrators will need to customize for their hub.

The most substantial of these, the *tasks* configuration file (see **Supplementary Data 1** for an example of the FluSight tasks configuration file), defines the modeling tasks to be addressed by a hub. To complete this configuration file, administrators will need to consider:

- the quantitative predictions to collect, or *modeling targets* (e.g., incident COVID-19 hospitalizations);
- the steps ahead being predicted, or *horizon* (e.g., 1-4 weeks in the future);
- other variables to be used in modeling, or *modeling task identifiers* (e.g., location);
- what dates define each round;
- what estimates to collect to represent the modeling targets, or *output type* (e.g., mean, median, quantile, probability mass function, etc.).

The hubAdmin R package^36^ can help create the tasks.json config file and can validate all configuration files.

Once configuration files are set up, hub administrators can accept model output submissions from modeling teams. The hub repository should contain a README file detailing the guidelines of the challenge to be addressed, including relevant dates, outcomes to be predicted, and data formatting requirements. Hub administrators create directories within the hub repository to collect both model output and *model metadata*. Model metadata describes the characteristics of models contributing to a hub, such as data inputs, methods, and how the model accounts for uncertainty. Model metadata may be used by hub administrators or data analysts to ensure that models are appropriate for the use case and for inclusion in the ensemble.

Hub administrators should ensure that all model output and metadata submissions are *validated*, i.e., tested against the configuration files to ensure the usability and integration of the output in downstream tools for data ingestion, ensembling, visualization, and evaluation. The `hubValidations` R package^37^ can be used to validate individual files or to set up ongoing validations of model output submitted to a hub repository. Hubs can also configure customized validation functions within standard `hubValidations` workflows if desired. A clearly defined tasks configuration file, together with the use of hubValidations, can help prevent mathematically invalid output and other problematic submissions.

Optionally, hub administrators may set up continuous integration through GitHub Actions to validate model submissions. Continuous integration involves automating the way code and data are validated prior to being merged into a shared hub repository. The hubverse currently provides several GitHub Action templates that can be used by GitHub-hosted hubs to validate model submissions and upload data to AWS S3 storage.

When predicted values of an outcome can be evaluated against actual observations, administrators should ensure that target data are available and accessible. Target data may need to be specifically formatted as *oracle output* data for use with evaluation tools (see Methods section for more on target data formats).

To communicate model output, administrators may create a simple website dashboard to provide model output visualizations and/or evaluation reports using hubverse dashboarding tools.

### Modelers

The active round phase of a hub revolves around modeling team submissions. Modelers are the key players in this phase and are responsible for submitting 1) *model output* data and 2) *model metadata* once for each model submitted to the hub repository, in conformance with submission guidelines. Modelers need to understand the different model output types that define the format in which quantitative predictions are structured and submitted to a hub (see Methods for more on model output). Both model output and model metadata can be validated before submission using the `hubValidations` R package.

### Data analysts

Data analysts perform secondary analyses of model output data in the post-round analysis phase of a hub. Data analysts may use hubverse tools to access model output, aggregate model output into ensembles, produce different model output visualizations, generate evaluation reports, or conduct secondary research.

The `hubData` R package^38^ leverages functionality from Apache Arrow^39^, allowing users to connect to, extract, query, and analyze model output data efficiently from the hub data store. Data analysts can connect to a fully configured hub or to the model output directory of a hub that has not been fully configured. In establishing a connection to a configured hub, the `hubData` package, using the R arrow package, creates a queryable dataset of files stored locally or in the cloud (analysts may also use standard tools for reading model output data, though this is less efficient).

The `hubEnsembles` package^40,41^ provides functionality to aggregate the output from multiple models into an ensemble using several common methodologies, such as Vincent ensembles and linear opinion pools^42^. The `hubEnsembles` package supports both unweighted (i.e., equally weighted) ensembles and weighted ensembles using unequal, user-specified weights for models. Data analysts may use different strategies to weight models included in ensembles^23–25^. For additional information on ensembling theory, methodology, and implementation with the hubEnsembles package, a tutorial-style manuscript is available^41^.

Analysts may also want to visualize and evaluate model output against target data. The `hubVis` R package^43^ contains functionality for plotting predictions that look at various time points in the future, along with optional target data. The `hubExamples` R package^44^ provides example model output and target data for an example forecast hub, and demonstrates how to join observed target values with model output to facilitate direct comparisons. The `hubEvals` R package^45^ contains a function to merge model output with target data and compute scores.

### Stakeholders

Stakeholders may interact with a hub through the consumption of hub products. We define three products of a hub that stakeholders are most likely to consult:

1. Ensemble models: aggregation of the output from several different models;
2. Website dashboard: a simple website with custom information pages, interactive visualizations, and/or model output evaluation;
3. Reports: customized model output material to share with different stakeholders.

### Developers

Developers offer overarching support that is not phase specific. Developers may help hub administrators set up new hubs and contribute new ideas, code, and documentation to the hubverse. Developers may work to implement new infrastructure (e.g., cloud storage architecture) and/or work on feature requests or bug fixes for existing hubverse software. The `hubExamples` package^44^ contains example model output of different output types that can be used to develop unit tests or document functions with code samples.

### Case study: U.S. CDC FluSight exercise

We use the FluSight Forecast Hub as a case study to demonstrate hubverse setup and functionality. The FluSight challenge, run annually by the U.S. CDC since 2013 (with a break for the 2021-2022 season due to reduced influenza activity during the COVID-19 pandemic), has served as a centrally coordinated collaborative effort to monitor and predict short-term influenza activity in the U.S. at the national, regional, and state level (https://github.com/cdcepi/FluSight-forecast-hub/)^26^. These seasonal collaborative forecasting challenges have garnered participation from dozens of academic, industry, and governmental research teams. Typically, forecasts have been submitted once a week from October through May. Forecasts consisted of quantitative predictions of observed values from public health surveillance systems that are consistently reported across the country.

During the 2023-2024 and 2024-2025 seasons, FluSight used the hubverse ecosystem to configure and maintain submissions of 1-4 week ahead forecasts (https://github.com/cdcepi/FluSight-forecast-hub/releases/tag/v1.0.0). **Supplementary Data 1** shows the tasks configuration file for the FluSight hub. In these seasons, teams could choose to submit forecasts for either or both of two modeling tasks:

TASK 1: Predicting the count of new weekly hospital admissions due to influenza

TASK 2: Predicting the category corresponding to the rate of change in hospital admissions (i.e., “large_decrease”, “decrease”, “stable”, “increase”, “large_increase”)

For both tasks, forecasts were accepted for 53 locations (50 states, the U.S., Puerto Rico, and Washington D.C.) and were made at the weekly scale. Forecasts were submitted every Wednesday during the season. Forecasts could be made for five different weekly prediction horizons (−1 through 3), where weeks were defined as Sunday through Saturday and corresponded to the definition of “MMWR weeks” used by the CDC^46^. A prediction made for a horizon of -1 referred to the prediction for the MMWR week that ended on the Saturday prior to the submission date (the previous week), and a prediction made for a horizon of 0 corresponded to the week ending on the Saturday after the Wednesday submission due date (the current week). Key properties of the two modeling tasks are summarized in **Table 2**.

**Table 2.** Information about the two modeling tasks for the U.S. FluSight challenge in both seasons.

| Name | Output type | Variable Type | Units |
| --- | --- | --- | --- |
| wk inc flu hosp | quantile | continuous | patient count |
| wk flu hosp rate change | probability mass function (pmf) | ordinal | categories based on rate per 100k population |

The “wk inc flu hosp” target represents the number of new laboratory-confirmed influenza incident hospital admissions in a given week. The units of this target are a patient count. The variable type is specified as “continuous” because, even though the target is an integer count of patients, the continuous specification allows models to represent prediction interval values as fractional values (a reasonable modeling choice) without data validation errors. The rate change target has five valid categories (“large_decrease”, “decrease”, “stable”, “increase”, “large_increase”) that can each be assigned a probability, where the five probabilities must sum to one. The precise mathematical definitions to resolve the category for a given location and week are described in the FluSight challenge guidelines (https://github.com/cdcepi/FluSight-forecast-hub/blob/v1.0.0/model-output/README.md#rate-trend-forecast-specifications). We note here only that the definitions are created such that weeks or locations with low counts where distinguishing between noise from increases and decreases can be difficult are resolved to the “stable” category.

A detailed code demonstration is provided in **Supplementary Note 1**.

## Discussion

The hubverse fills an important gap by providing a set of tools that standardizes predictive model output and stakeholder communication products. This open-source project provides infrastructure to 1) set up collaborative modeling hubs and collect model output submissions, and 2) serve as a central communication point for stakeholders.

In providing a standard infrastructure for collecting, aggregating, analyzing, and reporting model output, the hubverse eliminates duplication of effort in setting up code across different hubs. Usage of standard infrastructure lowers technological barriers to setting up a hub, which could allow for faster setup during emerging public health threats. For example, the U.S. COVID-19 Forecast Hub could likely have been set up even earlier in the pandemic if hubverse infrastructure had been available. Results of a recent Council of State and Territorial Epidemiologists survey sent out to the epidemiology workforce in the U.S. support the urgency of predictive modeling output during a pandemic: 45 out of 50 U.S. states and territories polled indicated that predictive modeling would be useful in their jurisdiction to inform decision-making during the next public health emergency^47^.

The hubverse software suite can be used to generate stakeholder communication products, including ensembles, interactive online dashboards, and evaluation reports. Ensemble models originated in weather forecasting^48^ but have become best practice in other fields, where they have been shown to result in improved accuracy^49,50^ and reliability^51^ and to produce more useful model output for decision-making^52^. Interactive online dashboards help stakeholders visualize model output and uncertainty, which is increasingly important in order to build trust^4^. Evaluation reports provide evidence of model performance that can be used by decision-makers to select or discard models for use in decision-making^53^.

In the field of infectious disease, hubverse-style hubs have been used to generate projections that respond to different public health questions. Forecasts are quantitative predictions of future disease trends, and can help answer questions about what is likely to happen in the near-term future. Forecasts can help guide decisions on operational planning and the amount of intervention needed to control disease spread. Nowcasts provide predictions about the current state of an outbreak by adjusting for data lags from a data stream up until the current date, helping to answer questions about what is happening now. They provide information about the current situation and can help guide strategies to manage immediate needs. Both nowcasts and forecasts can be evaluated against target data. In contrast, scenario projections are predictions that answer “what if” questions about disease trends if certain assumptions are met (e.g., disease transmissibility, vaccine efficacy or uptake, interventions, emergence of new variants). In contrast to forecasts and nowcasts, it is difficult to evaluate scenario projections since the assumptions these predictions are based upon are unlikely to ever be completely met. However, scenario projections can be useful tools to evaluate possible longer-term (e.g. spanning many months to years) patterns of transmission or the effectiveness of interventions, and may thus offer information for longer-term strategic planning^18,54^. Importantly, there are no technical barriers in using hubverse infrastructure to collect forecasts at extended time horizons. Additionally, hubverse-style hubs can also be used to aggregate estimates of disease parameters (e.g., incubation period, transmission rate, waning of immunity rate). Parameter estimation may be useful on its own to better understand epidemiological characteristics that can guide prevention and control measures or these results could be incorporated into models to improve model performance.

While hubverse infrastructure is flexible enough to support applications across domains, its adoption is shaped by the availability of financial, computational, and organizational resources. Although hubverse does not remove the need for these resources, it can lower entry barriers by reducing operational overhead and leveraging no-cost infrastructure. All hubverse tools are open source and hosting services, such as GitHub, are free to use. Additionally, in lower-resource settings where there are limited model contributors, independently-developed tools aligned with hub-style workflows have emerged. For example, the open-source MicroHub tool^55^ has been used to support standardized forecast generation in Paraguay.

The current hubverse model output data format is tied to a tabular data structure that can be inefficient for larger datasets, thus limiting the complexity of modeling scenarios and/or hub setup. Currently, large hubs can use Parquet, a binary file format that stores data by column, resulting in more efficient data compression and faster data querying^56^. Additionally, users can partition Parquet files by common query dimensions (e.g., date, location, or target), which would allow tools like hubData to load only the needed subsets, though this is currently only available for target data. Another limitation is that data-type specifications for the “value” column can be challenging when some tasks have outputs that require strings while others require numeric values (e.g., one task might involve a label such as “decrease”, while another might have a numeric value). Furthermore, hubverse tools are dependent on use with specific file formats (CSV, Parquet) and are mainly available with R, though some Python development is in progress. Lastly, the current infrastructure is only available in English, though there is capacity to accommodate other languages.

The are four large-scale upcoming projects on the hubverse roadmap. The first allows for the creation of cloud-based hubs that can accept submissions directly, without needing to be mirrored to a repository-based hub. Cloud-based hubs would permit larger file sizes, allowing for the collection of more samples and modeling tasks, including a breakdown into smaller subpopulations. The second involves containerizing models to run on cloud infrastructure. This would allow for a hub to come with a few standard models that could be run on data provided by hub administrators, with a standard output to interface with the hubverse. The third would create “benchmark hubs” to serve as research-ready repositories for machine-learning researchers looking to test new methods and compare them with existing models. These might include creating an archival hubverse version of several of the large modeling hubs that were run prior to the existence of hubverse standards. The fourth involves conducting retrospective evaluations of hub model output, as made possible by hubverse standards and archiving.

In summary, as increasing attention is paid to predictive modeling, the hubverse provides infrastructure to improve the scientific rigor behind real-time and retrospective predictive modeling challenges.

## Methods

### File storage architecture

Hubverse-style hubs maintain a file-based data storage architecture. Hubs should contain the directories, subdirectories, and files shown in **Extended Data Table 2**. In brief, all hubs should have a documentation file (such as a README file) at the top level containing information on the hub structure. All hubs must also have a hub configuration directory containing all configuration files. Hubs will have separate directories for model output and model metadata submissions from modeling teams. Hubs that predict outcomes that may be eventually evaluated against target data may store these data within an optional target data directory. If any code and scripts are present in a hub repository, they should be stored within the source code directory, and never within the model output directory.

While other options for data storage exist, namely databases, the decision to organize hub data storage around files was based on several pragmatic considerations. First, files are the natural submission unit for hubs. Using the original files allows a hub to take advantage of existing version-control software with time-stamps for clear audit trails of when files were submitted, as well as setting up clear “per-file” continuous integration actions, such as data validation or transformation. Second, when stored in an Arrow-compliant file-based structure, data at the scale of most current hubs can be retrieved with similar speed as if it were in a lightweight database (the hubverse development team ran some benchmarking experiments comparing a DuckDB to a specific Arrow-based partition of the files). Third, keeping a file-structure-based storage system lowers barriers to entry for groups that want to set up a hub and leverage hubverse tools but don’t have the capacity or technical expertise to stand up a database backend.

### Model output standards

Modeling team submissions are known as model output. Model output must follow a tabular representation where each row represents a single prediction (or one aspect of a prediction, like a single quantile value, or a single sample) and each column provides additional information about the prediction. Model output may be submitted as CSV or Parquet.

Model output columns can be divided into four groups: (1) the *model identifier* indicates what model has produced the prediction, (2) the *task identifiers (task IDs)* provide details about what is being predicted, (3) *output representation* specifies how the prediction is reported (e.g., mean, median, quantile), and (4) *value* provides the prediction. The latter three groups must correspond to how modeling tasks were defined in the tasks.json configuration file (see **Supplementary Data 1** for example tasks configuration from the FluSight hub). Example model output displaying model identifier, task identifier, output representation, and value columns are shown in **Extended Data Table 3**.

*Task IDs:* Task IDs define the variables included in the model output and how they should be represented. Each unique row of model output data is defined by a combination of task ID values and output representation specifications (output_type and output_type_id). The composition of the task ID variables and their accepted values are hub specific. Commonly used (and optional) task IDs are shown in **Extended Data Table 4,** but additional task IDs may be specified depending on the needs of a hub. The hub can specify the structure, data type, and valid values of all task ID variables.

Hubs often use a task ID variable to define a submission round, which may be as simple as using origin_date or forecast_date.

*Output representation:* This component defines how the prediction is represented in model output submissions. The output_type column defines the type of representation of the predictive distribution, while the output_type_id provides additional identifying information specific to the output type. These two columns can be thought of as providing output metadata about the predicted value.

The *value* column provides the prediction.

See **Table 3** for output and value representations.

**Table 3.**
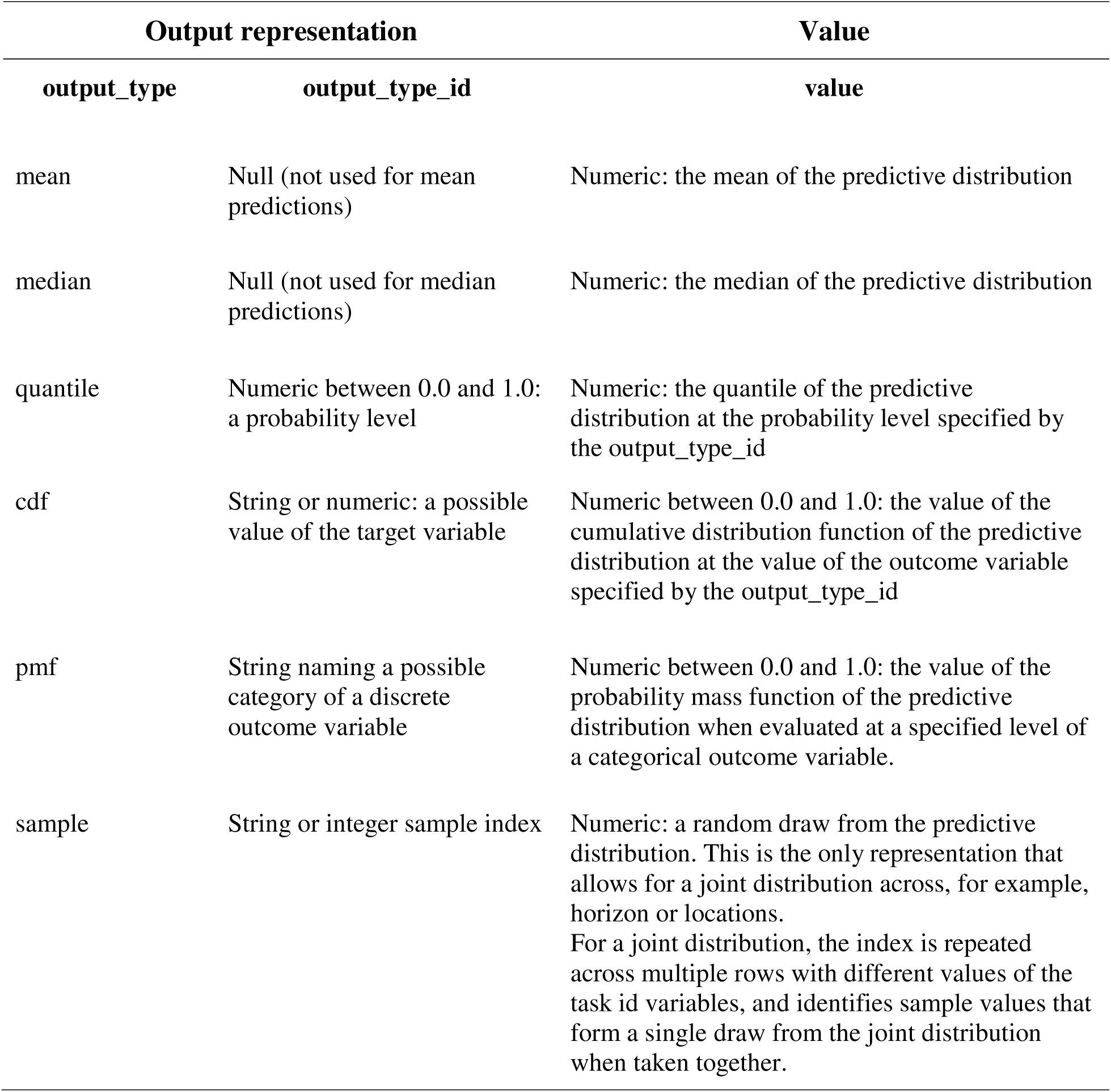
Output and value representations.

### Target data

Target data are the actual observed values of what is being predicted in model submissions. Hubs that evaluate model predictions against target data values should ensure that target data are available and accessible. This can most commonly be achieved by providing code to access target data or by storing target data snapshots in the target data directory within the hub repository. Target data can be represented in two forms: *time-series* or *oracle output*.

The first format is *time-series* data. This is often the native or “raw” format for data. Each row of the dataset contains one unit of observation. For example, if the number of influenza cases per week is being reported for each of several states, the unit of observation would be a location and week. The columns consist of:

1. Task ID variables that uniquely define the unit of observation, with one column representing the date.
2. An “observation” column with the observed value.

The second format is *oracle output* data. Oracle output data are derived from the time series data and represents model output that would have been generated if the target data values had been known in advance, as if by an oracle. Oracle output follows a format similar to a hubverse model output file, with three main differences:

1. Predictions correspond to a distribution that places probability 1 on the observed target outcome.
2. Predictions are stored in a column named ‘oracle_valuè rather than ’valuè. The implications of this depend on the output_type (**Extended Data Table 5**).
3. Generally, oracle output columns will be a subset of the columns of valid model output, with just those columns that are needed to correctly align oracle_values with the corresponding model output predicted values.

This structure allows a model output dataset to be joined with an oracle output dataset so that the model output ‘valuè can be compared and evaluated against the corresponding ’oracle_valuè. Having these data together in the same row can be helpful for evaluation and visualization. **Table 4** shows an example of specially formatted oracle output data, which can then be joined to model output data for visualization and evaluation purposes.

**Table 4.** Example oracle output data, which can be joined to model output data for visualization and evaluation purposes. Note that oracle output data will contain the subset of task identifier columns needed to align model output and oracle output data.

| Oracle output data |  |  |  |  |  |
| --- | --- | --- | --- | --- | --- |
| Task identifiers |  |  | Output representation |  | Oracle value |
| location | target_end_date | target | output_type | output_type_id | oracle_value |
| **\*mean** |  |  |  |  |  |
| 25 | 2022-11-19 | wk inc flu hosp | mean | NA | 79 |
| **\*median** |  |  |  |  |  |
| 25 | 2022-11-19 | wk inc flu hosp | median | NA | 79 |
| **\*quantile** |  |  |  |  |  |
| 25 | 2022-11-19 | wk inc flu hosp | quantile | NA | 79 |
| **\*pmf** |  |  |  |  |  |
| 25 | 2022-11-19 | wk flu hosp rate category | pmf | low | 1 |
| 25 | 2022-11-19 | wk flu hosp rate category | pmf | moderate | 0 |
| 25 | 2022-11-19 | wk flu hosp rate category | pmf | high | 0 |
| 25 | 2022-11-19 | wk flu hosp rate category | pmf | very high | 0 |
| **\*cdf** |  |  |  |  |  |
| 25 | 2022-11-19 | wk flu hosp rate | cdf | 0.75 | 0 |
| 25 | 2022-11-19 | wk flu hosp rate | cdf | 1 | 0 |
| 25 | 2022-11-19 | wk flu hosp rate | cdf | 1.25 | 1 |
| 25 | 2022-11-20 | wk flu hosp rate | cdf | 1.5 | 1 |
| **\*sample** |  |  |  |  |  |
| 25 | 2022-11-19 | wk inc flu hosp | sample | NA | 79 |

\*These rows label the output type for readability only and are not part of a standard oracle output file

### Technology and data formats used in development

All hubverse code is developed in public, open source repositories on Github. Hubverse data may live on GitHub and/or on cloud servers, such as AWS S3 buckets. The R^57^ and Python^58^ languages are used to develop user-facing libraries. The Apache Arrow^39^ columnar data format is used when data are read in by users.

Datafiles may be stored in CSV or Parquet formats. JSON format textfiles are used for hub configuration files and JSON or YAML files are accepted for model metadata. Cloud storage for hubverse repositories are set up using infrastructure-as-code services such as Pulumi. Continuous integration, such as used for real-time validation of submitted model output, is implemented via GitHub Actions.

### Contributions to FAIR

The hubverse promotes interoperability, a core component of FAIR (findable, accessible, interoperable, re-usable) principles, through the use of standardized schema, validation infrastructure, and common data formats that facilitate data synthesis and ensemble models. The current infrastructure is openly accessible in practice, though it does not yet meet formal accessibility requirements. Strengthening findability and re-usability through DOIs, registration in searchable repositories, and structured metadata schemas remain important areas for future development.

## Consortium Authors

The following authors are members of the Consortium of Infectious Disease Modeling Hubs and are included in the main author list of this manuscript:

## Data Availability

Datasets showing example model output and oracle output data are available in the hubExamples R package, which provides example data for forecasting and scenario modeling hubs in the hubverse format, https://github.com/hubverse-org/hubExamples.

U.S. CDC FluSight model output and target data datasets from the 2023-2024 flu season are available on GitHub: https://github.com/cdcepi/FluSight-forecast-hub/releases/tag/v1.0.0. The tasks configuration file is also available at this link and in the Supplementary Data.

Hubs with publicly available datasets and S3 buckets are available here: https://hubverse.io/community/hubs.html

## Code Availability

All code related to the hubverse project is publicly available under an MIT License. The code underlying this study is publicly available through the links provided and versioned, with a release corresponding to the analyses presented in the case study (see last item). The repository is maintained by the authors and will be updated as part of ongoing research activities, contingent on available funding. We aim to address critical issues and bug fixes as feasible, but do not guarantee continuous long-term maintenance. External contributions may be submitted via pull requests and will be reviewed by the maintainers.

No new datasets were generated for the overview component of this manuscript. All tools discussed are publicly available, and links to their current versions are provided in the manuscript (accessed January–April 2026).

Code to create and validate hub configuration files is available in the hubAdmin R package: https://github.com/hubverse-org/hubAdmin.

Code to validate model output and model metadata submissions is available in the hubValidations R package: https://github.com/hubverse-org/hubValidations.

Code to set up continuous integration is available in the hubCI R package: https://github.com/hubverse-org/hubCI.

Code to connect to hub data is available in the hubData R package: https://github.com/hubverse-org/hubData.

Code to aggregate model output into ensembles is available in the hubEnsembles R package: https://github.com/hubverse-org/hubEnsembles.

Code to visualize model output is available in the hubVis R package: https://github.com/hubverse-org/hubVis.

Code to join observed target data values with model output in order to facilitate direct comparisons is available in the hubExamples R package: https://github.com/hubverse-org/hubExamples.

Code to evaluate model output is available in the hubEvals R package: https://github.com/hubverse-org/hubEvals.

Code related to all packages listed above is also available in the hubverse R package: https://github.com/hubverse-org/hubverse.

The computational environment for the FluSight case study, including R version, operating system, and full package version information, is provided in the *Session information* section of the vignette included in the Supplementary Information and available at https://github.com/hubverse-org/hubverse/blob/main/vignettes/hubverse_ms_vignette.pdf.

## Disclaimer

The findings and conclusions in this report are those of the authors and do not necessarily represent the official position of the Centers for Disease Control and Prevention. The content of this publication does not necessarily reflect the views or policies of the Department of Health and Human Services, nor does mention of trade names, commercial products or organizations imply endorsement by the U.S. Government.

## Data Availability

Data Availability
Datasets showing example model output and oracle output data are available in the hubExamples R package, which provides example data for forecasting and scenario modeling hubs in the hubverse format, https://github.com/hubverse-org/hubExamples.
U.S. CDC FluSight model output and target data datasets from the 2023-2024 flu season are available on GitHub: https://github.com/cdcepi/FluSight-forecast-hub/releases/tag/v1.0.0.
Hubs with publicly available datasets and S3 buckets are available here: https://hubverse.io/community/hubs.html
Code Availability
All code related to the hubverse project is publicly available under an MIT License.
Code to create and validate hub configuration files is available in the hubAdmin R package: https://github.com/hubverse-org/hubAdmin.
Code to validate model output and model metadata submissions is available in the hubValidations R package: https://github.com/hubverse-org/hubValidations.
Code to set up continuous integration is available in the hubCI R package: https://github.com/hubverse-org/hubCI.
Code to connect to hub data is available in the hubData R package: https://github.com/hubverse-org/hubData.
Code to aggregate model output into ensembles is available in the hubEnsembles R package: https://github.com/hubverse-org/hubEnsembles.
Code to visualize model output is available in the hubVis R package: https://github.com/hubverse-org/hubVis.
Code to join observed target data values with model output in order to facilitate direct comparisons is available in the hubExamples R package: https://github.com/hubverse-org/hubExamples.
Code to evaluate model output is available in the hubEvals R package: https://github.com/hubverse-org/hubEvals.
Code related to all packages listed above is also available in the hubverse R package: https://github.com/hubverse-org/hubverse.

## Acknowledgements

The authors would like to thank Zhian Kamvar for the figure presenting an overview of hub architecture and Rebecca Sweger for the figure presenting a schematic of the hubverse showing different user roles.

## Author Contributions

Melissa Kerr: Writing – review & editing, Writing – original draft, Conceptualization. Rebecca Borchering: Review & editing. Alvaro Castro Rivadeneira: Review & editing. Lucie Contamin: Review & editing. Sebastian Funk: Review & editing. Harry Hochheiser: Review & editing. Emily Howerton: Review & editing. Anna Krystalli: Review & editing. Li Shandross: Review & editing. Nicholas G. Reich: Writing – review & editing, Writing – original draft, Conceptualization, Supervision.

## Competing Interests

N.G.R. discloses independent paid consulting with Google Research. The remaining authors declare no competing interests.

| Hub | Owner | Year(s) | Repository (https://github.com/) | Rows of model output data (as of June 2025) | # Models | Hubverse-developed dashboard | Cloud storage |
| --- | --- | --- | --- | --- | --- | --- | --- |
| **Community hubs** |  |  |  |  |  |  |  |
| COVID-19 Scenario Modeling Hub* | Scenario Modeling Hub Coordination Team | * | [midas-network/covid19-scenario-modeling-hub](https://github.com/midas-network/covid19-scenario-modeling-hub) | 626,219,176 | 28 |  |  |
| RespiCompass | European Centre for Disease Prevention and Control (ECDC) | 2024- | [european-modelling-hubs/RespiCompass](https://github.com/european-modelling-hubs/RespiCompass) | 213,562,080 | 14 |  |  |
| SARS-CoV-2 Variant Nowcast Hub | Reich Lab at UMass-Amherst | 2024- | [reichlab/variant-nowcast-hub](https://github.com/reichlab/variant-nowcast-hub) | 204,133,734 | 6 | X | X |
| Flu Scenario Modeling Hub* | Scenario Modeling Hub Coordination Team | * | [midas-network/flu-scenario-modeling-hub](https://github.com/midas-network/flu-scenario-modeling-hub) | 150,013,626 | 19 |  |  |
| RSV Scenario Modeling Hub* | Scenario Modeling Hub Coordination Team | * | [midas-network/rsv-scenario-modeling-hub](https://github.com/midas-network/rsv-scenario-modeling-hub) | 55,188,380 | 18 |  |  |
| FluSight archive | Hubverse | 2015-2020 | [hubverse-org/flusight_hub_archive](https://github.com/hubverse-org/flusight_hub_archive) | 29,364,474 | 99 |  | X |
| FluSight | U.S. Centers for Disease Control and Prevention (CDC) | 2023- | [cdcepi/FluSight-forecast-hub/](https://github.com/cdcepi/FluSight-forecast-hub/) | 13,832,587 | 70 | X | X |
| COVID-19 Scenario Modeling Hub - Research* | Scenario Modeling Hub Coordination Team | * | [midas-network/covid19-smh-research/](https://github.com/midas-network/covid19-smh-research/) | 3,185,180 | 7 |  |  |
| COVID-19 Forecast Hub | CDC | 2024- | [CDCgov/covid19-forecast-hub](https://github.com/CDCgov/covid19-forecast-hub) | 1,993,088 | 19 | X | X |
| RespiCast Syndromic Indicators | ECDC | 2024- | [european-modelling-hubs/RespiCast-SyndromicIndicators](https://github.com/european-modelling-hubs/RespiCast-SyndromicIndicators) | 1,591,830 | 23 |  |  |
| US RSV Forecast Hub | Johns Hopkins University Infectious Disease Dynamics Group | 2023- | [HopkinsIDD/rsv-forecast-hub](https://github.com/HopkinsIDD/rsv-forecast-hub) | 1,117,639 | 9 |  |  |
| RespiCast Influenza archive | ECDC | 2023-2024 | [european-modelling-hubs/flu-forecast-hub_archive](https://github.com/european-modelling-hubs/flu-forecast-hub_archive) | 407,938 | 16 |  |  |
| RespiCast COVID-19 | ECDC | 2024- | [european-modelling-hubs/RespiCast-Covid19](https://github.com/european-modelling-hubs/RespiCast-Covid19) | 358,931 | 13 |  |  |
| AI4Casting Hub Respiratory Virus | University of Guelph | 2024- | [ai4castinghub/rvdss-forecast](https://github.com/ai4castinghub/rvdss-forecast) | 255,640 | 9 |  |  |
| RespiCast ARI archive | ECDC | 2023-2024 | [european-modelling-hubs/ari-forecast-hub_archive](https://github.com/european-modelling-hubs/ari-forecast-hub_archive) | 218,844 | 12 |  |  |
| AI4Casting Hub Hospital Bed Occupancy | University of Guelph | 2024- | [ai4castinghub/hospitalization-forecast](https://github.com/ai4castinghub/hospitalization-forecast) | 90,055 | 6 |  |  |
| West Nile Virus Forecast Hub | California Department of Public Health | 2024- | [cdphmodeling/wnvca-2024](https://github.com/cdphmodeling/wnvca-2024) | 61,061 | 25 |  |  |
| Flu Metrocast Hub | University of Texas-Austin and UMass-Amherst | 2025- | [reichlab/flu-metrocast](https://github.com/reichlab/flu-metrocast) | 49,968 | 7 | X | X |
| **Collaborative hubs with private repositories** |  |  |  |  |  |  |  |
| North Carolina Department of Health and Human Services/Atlantic Coast Center for Infectious Disease Dynamics and Analytics (ACCIDDA) Forecasting Collaboration | ACCIDDA |  | Private | Private | Private |  |  |
| Paraguay Respiratory Virus Forecast Hub | University of Georgia/US CDC |  | Private | Private | Private |  |  |
| Australia-Aotearoa Forecasting Hub | Australia-Aotearoa Consortium for Epidemic Forecasting and Analytics |  | Private | Private | Private |  |  |
| **Local hubs** |  |  |  |  |  |  |  |
| Variant Nowcast Model Development Retrospective Hub | Reich Lab at UMass-Amherst |  | https://github.com/reichlab/variant-nowcast-model- | 1,291,807,776 | 8 |  |  |
| Flusion- Retrospective Hub | Reich Lab at UMass-Amherst |  | https://github.com/reichlab/flusion/tree/main/retros | 1,376,251 | 9 |  |  |
| Flusion- Submissions Hub | Reich Lab at UMass-Amherst |  | https://github.com/reichlab/flusion/tree/main/submi | 765,440 | 8 |  |  |

\* Pre-existing hub transitioning to hubverse standards and tools.
† Based on model output data contained in the hub repository as of Sept. 2025.

Hub Structure
| Component | Name, location, and format within a hub | Description | Provided by: |  |
| --- | --- | --- | --- | --- |
| Documentation |  |  | Hub | Modeler |
| Documentation file | e.g., README.md file, located in top level of hub and within each directory | File containing info about the hub structure and additional details about each of the directories | ☒ |  |
| Hub configuration |  |  |  |  |
| Hub configuration directory | /hub-config/ | Folder storing configuration files | ☒ |  |
| Hub admin configuration file | /hub-config/admin.json | Structured text file containing overall configuration settings for the hub | ☒ |  |
| Hub modeling tasks configuration file | /hub-config/tasks.json | Structured text file that defines modeling tasks and therefore implicitly defines the assumed structure for any model submitted | ☒ |  |
| Hub model metadata configuration file | /hub-config/model-metadata-schema.json | Structured text file that defines the expected format of model metadata files submitted by modeling teams | ☒ |  |
| Modeling team submissions |  |  |  |  |
| Model output directory | /model-output/ | Folder to collect modeling team model submissions | ☒ |  |
| Model output subdirectory | /model-output/team1-modela | Model-specific subdirectory for submissions from one modeling team. |  | ☒ |
| Model output file | /model-output/team1-modela/<round-id1><model_id>.csv or .parquet | Round-specific model submission file |  | ☒ |
| Model metadata |  |  |  |  |
| Model metadata directory | /model-metadata/ | Folder to collect modeling team model metadata submissions | ☒ |  |
| Model metadata file | /model-metadata/team1-modela.yml or yaml | Model-specific metadata submission file |  | ☒ |
| Model abstracts (optional) |  |  |  |  |
| Model abstracts directory | /model-abstracts/ | Folder to collect optional round-specific model metadata | ☒ |  |
| Model abstract subdirectory | /model-abstracts/team1-modela/ | Model-specific subdirectory for round-specific model metadata |  | ☒ |
| Model abstract submission file | /model-abstracts/team1-modela/<round-id1>.md | Round-specific model metadata submission |  | ☒ |
| Target data directory (optional) |  |  |  |  |
| Target data directory | /target-data/ | Folder storing actual observed (i.e., target) values of an outcome, links to external open-access sources, and/or information on how model targets can be calculated from target data | ☒ |  |
| Time series data | /target-data/time-series.csv or .parquet | File with observed counts or rates | ☒ |  |
| Oracle output data | /target-data/oracle-output.csv or .parquet | File containing data dervied from time series data; represents the model output that would have been generated if the target data values were known ahead of time. | ☒ |  |
| Auxiliary data directory (optional) |  |  |  |  |
| Auxiliary data directory | /auxiliary-data/ | Folder to store any additional data related to modeling efforts | ☒ |  |
| Source code directory (optional) |  |  |  |  |
| Source code directory | /src/ | Folder storing code that is present within the hub repository | ☒ |  |

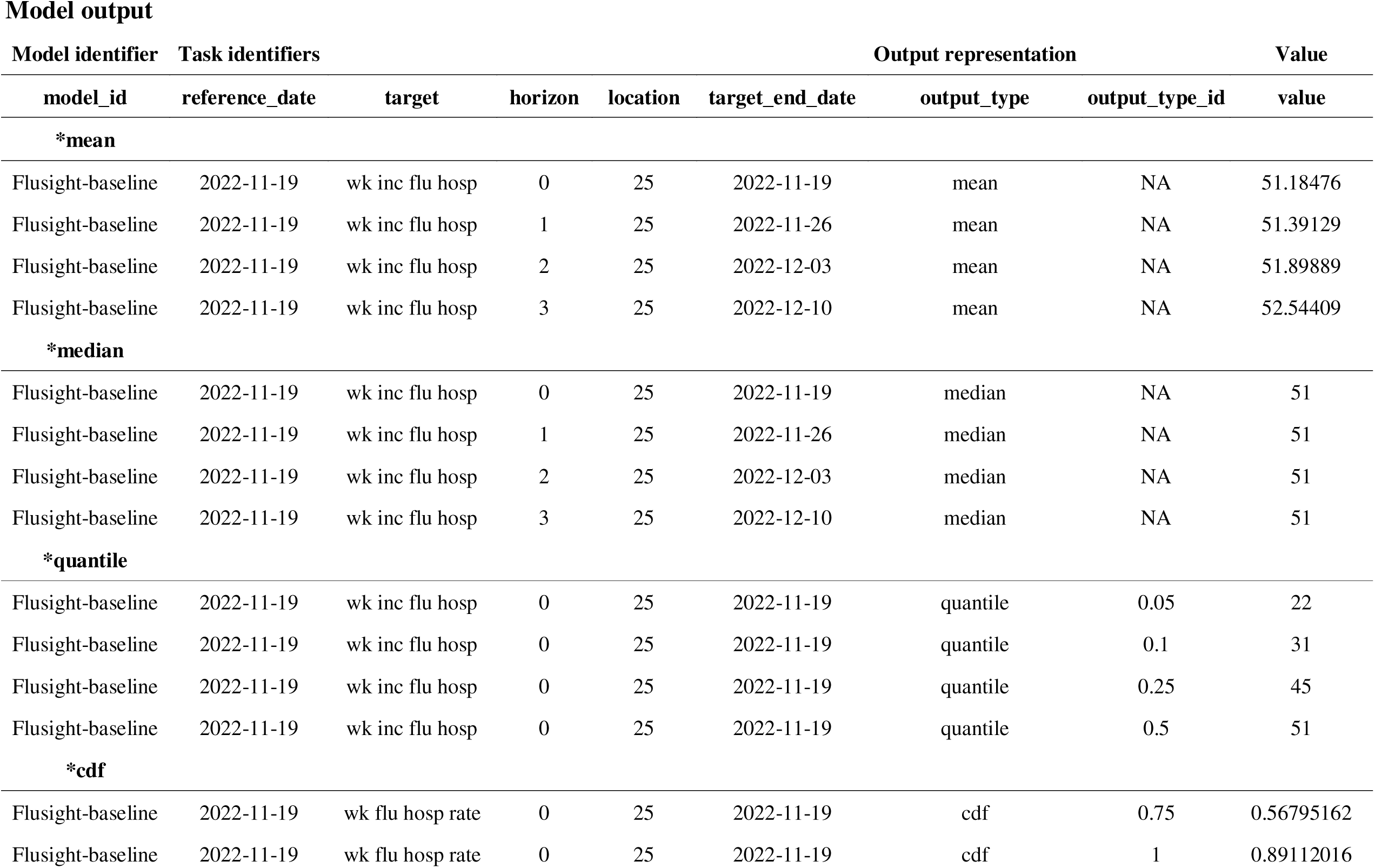
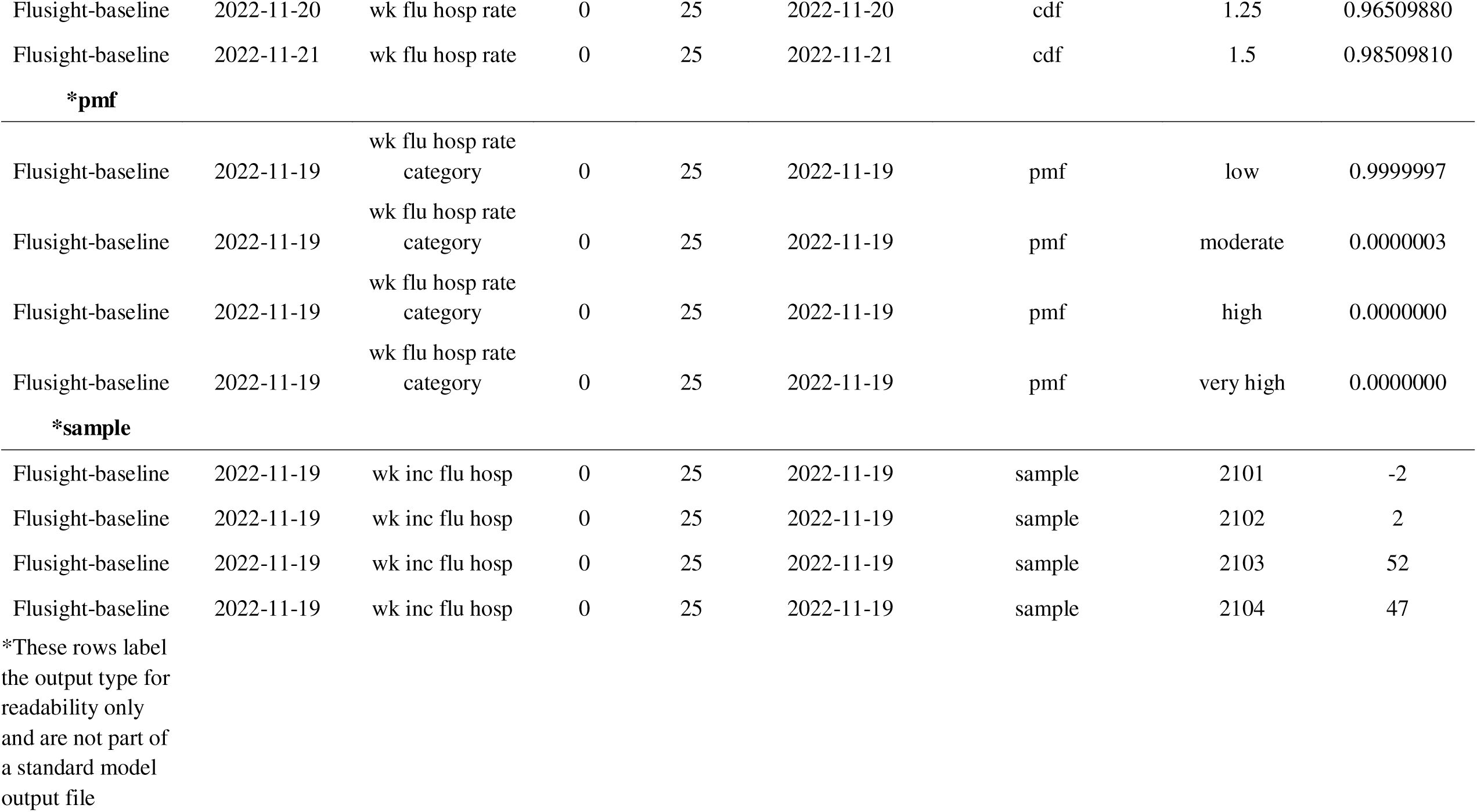

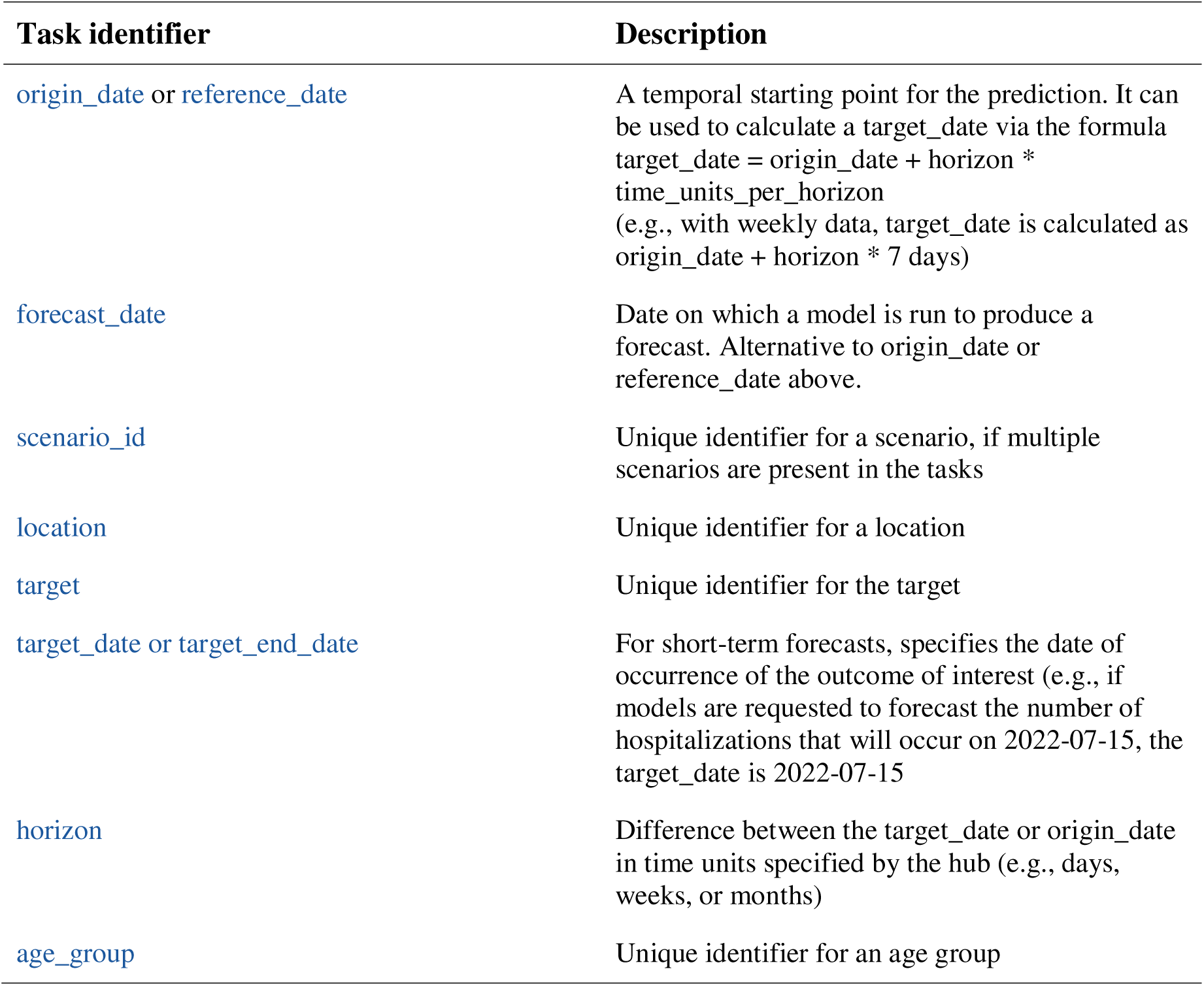
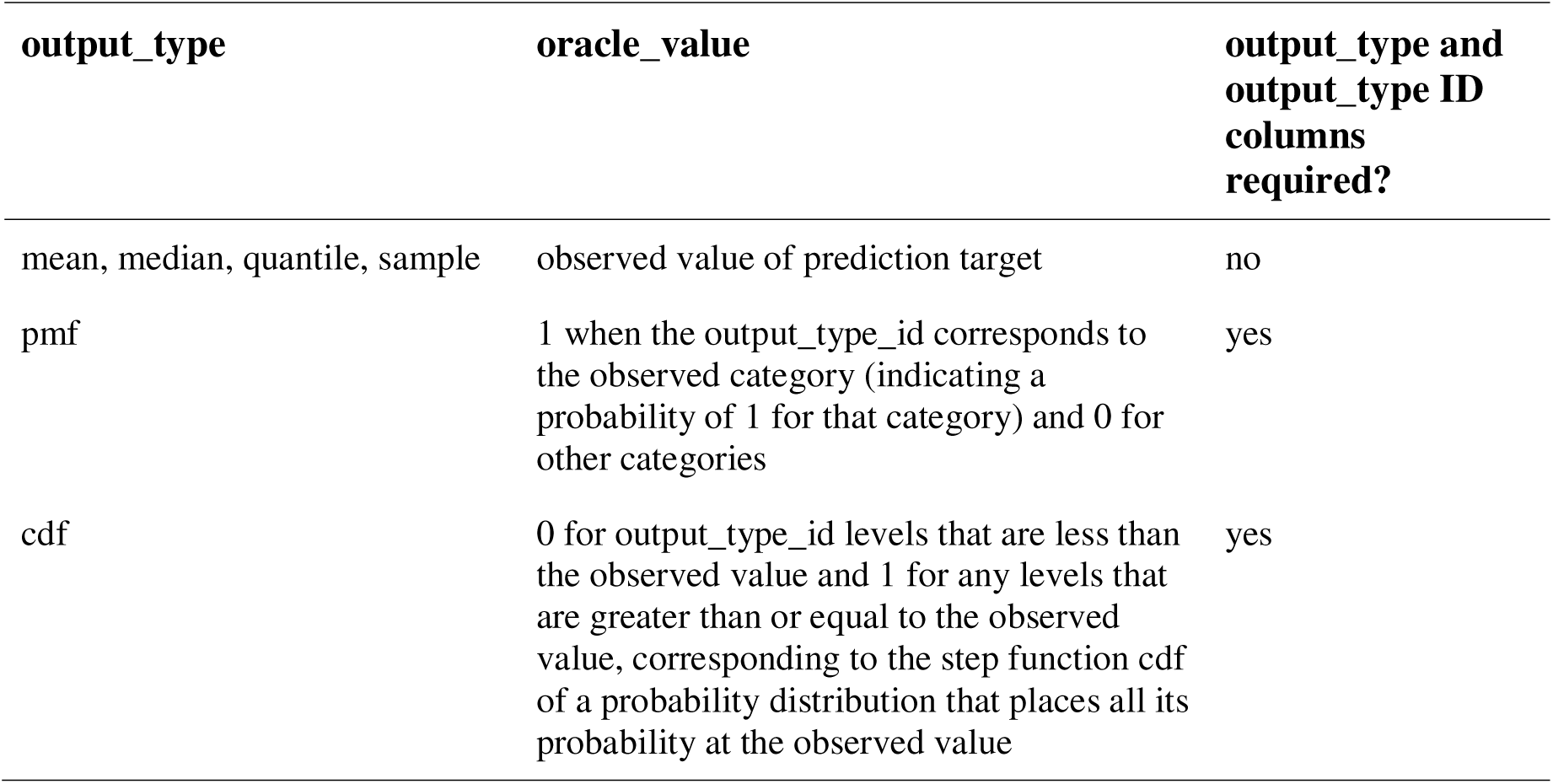

## References

1. Meltzer, M. I. et al. Modeling in Real Time During the Ebola Response. MMWR Suppl. 65, 85–89 (2016), doi:10.15585/mmwr.su6503a12.

2. Yang, H. et al. Design of COVID-19 staged alert systems to ensure healthcare capacity with minimal closures. Nat. Commun. 12, 3767 (2021), doi:10.1038/s41467-021-23989-x.

3. Borchering, R. K. et al. Public health impact of the U.S. Scenario Modeling Hub. Epidemics 44, 100705 (2023), doi:10.1016/j.epidem.2023.100705.

4. Bodner, K. et al. Bridging the divide between ecological forecasts and environmental decision making. Ecosphere 12, e03869 (2021), doi:10.1002/ecs2.3869.

5. Grimm, V., Johnston, A. S. A., Thulke, H.-H., Forbes, V. E. & Thorbek, P. Three questions to ask before using model outputs for decision support. Nat. Commun. 11, 4959 (2020), doi:10.1038/s41467-020-17785-2.

6. Chretien, J.-P., George, D., Shaman, J., Chitale, R. A. & McKenzie, F. E. Influenza forecasting in human populations: a scoping review. PloS One 9, e94130 (2014), doi:10.1371/journal.pone.0094130.

7. Li, X. et al. Estimating the health impact of vaccination against ten pathogens in 98 low-income and middle-income countries from 2000 to 2030: a modelling study. The Lancet 397, 398–408 (2021), doi:10.1016/S0140-6736(20)32657-X.

8. Houben, R. M. G. J. et al. Feasibility of achieving the 2025 WHO global tuberculosis targets in South Africa, China, and India: a combined analysis of 11 mathematical models. Lancet Glob. Health 4, e806–e815 (2016), doi:10.1016/S2214-109X(16)30199-1.

9. Brady, O. J. et al. Role of mass drug administration in elimination of Plasmodium falciparum malaria: a consensus modelling study. Lancet Glob. Health 5, e680–e687 (2017), doi:10.1016/S2214-109X(17)30220-6.

10. Flasche, S. et al. The Long-Term Safety, Public Health Impact, and Cost-Effectiveness of Routine Vaccination with a Recombinant, Live-Attenuated Dengue Vaccine (Dengvaxia): A Model Comparison Study. PLOS Med. 13, e1002181 (2016), doi:10.1371/journal.pmed.1002181.

11. Jung, S.-M. et al. Potential impact of annual vaccination with reformulated COVID-19 vaccines: Lessons from the US COVID-19 scenario modeling hub. PLoS Med. 21, e1004387 (2024), doi:10.1371/journal.pmed.1004387.

12. Loo, S. L. et al. The US COVID-19 and Influenza Scenario Modeling Hubs: Delivering long-term projections to guide policy. Epidemics 46, 100738 (2024), doi:10.1016/j.epidem.2023.100738.

13. Biggerstaff, M. et al. Results from the centers for disease control and prevention’s predict the 2013–2014 Influenza Season Challenge. BMC Infect. Dis. 16, 357 (2016), doi:10.1186/s12879-016-1669-x.

14. Cramer, E. Y., et al. The United States COVID-19 Forecast Hub dataset. Sci. Data 9, 462 (2022), doi:10.1038/s41597-022-01517-w.

15. Johansson, M. A. et al. An open challenge to advance probabilistic forecasting for dengue epidemics. Proc. Natl. Acad. Sci. U. S. A. 116, 24268–24274 (2019), doi:10.1073/pnas.1909865116.

16. Viboud, C. et al. The RAPIDD ebola forecasting challenge: Synthesis and lessons learnt. Epidemics 22, 13–21 (2018), doi:10.1016/j.epidem.2017.08.002.

17. Shea, K. et al. Multiple models for outbreak decision support in the face of uncertainty. Proc. Natl. Acad. Sci. 120, e2207537120 (2023), doi:10.1073/pnas.2207537120.

18. Reich, N. G. et al. Collaborative Hubs: Making the Most of Predictive Epidemic Modeling. Am. J. Public Health 112, 839–842 (2022), doi:10.2105/AJPH.2022.306831.

19. Pennell, C. & Reichler, T. On the Effective Number of Climate Models. (2011) doi:10.1175/2010JCLI3814.1, doi:10.1175/2010JCLI3814.1.

20. Reich, N. G. et al. Accuracy of real-time multi-model ensemble forecasts for seasonal influenza in the U.S. PLoS Comput. Biol. 15, e1007486 (2019), doi:10.1371/journal.pcbi.1007486.

21. Cramer, E. Y. et al. Evaluation of individual and ensemble probabilistic forecasts of COVID-19 mortality in the United States. Proc. Natl. Acad. Sci. 119, e2113561119 (2022), doi:10.1073/pnas.2113561119.

22. Sherratt, K. et al. Predictive performance of multi-model ensemble forecasts of COVID-19 across European nations. eLife 12, e81916 (2023), doi:10.7554/eLife.81916.

23. Ray, E. L. & Reich, N. G. Prediction of infectious disease epidemics via weighted density ensembles. PLoS Comput. Biol. 14, e1005910 (2018), doi:10.1371/journal.pcbi.1005910.

24. Oidtman, R. J. et al. Trade-offs between individual and ensemble forecasts of an emerging infectious disease. Nat. Commun. 12, 5379 (2021), doi:10.1038/s41467-021-25695-0.

25. McAndrew, T. & Reich, N. G. Adaptively stacking ensembles for influenza forecasting. Stat. Med. 40, 6931–6952 (2021), doi:10.1002/sim.9219.

26. Lutz, C. S. et al. Applying infectious disease forecasting to public health: a path forward using influenza forecasting examples. BMC Public Health 19, 1659 (2019), doi:10.1186/s12889-019-7966-8.

27. Reich, N. G. et al. A collaborative multiyear, multimodel assessment of seasonal influenza forecasting in the United States. Proc. Natl. Acad. Sci. U. S. A. 116, 3146–3154 (2019), doi:10.1073/pnas.1812594116.

28. Borchering, R. K. et al. Impact of SARS-CoV-2 vaccination of children ages 5–11 years on COVID-19 disease burden and resilience to new variants in the United States, November 2021–March 2022: a multi-model study. Lancet Reg. Health - Am. 17, 100398 (2022), doi:10.1016/j.lana.2022.100398.

29. Rosenblum, H. G. et al. Interim Recommendations from the Advisory Committee on Immunization Practices for the Use of Bivalent Booster Doses of COVID-19 Vaccines — United States, October 2022. Morb. Mortal. Wkly. Rep. 71, 1436–1441 (2022), doi:10.15585/mmwr.mm7145a2.

30. Mathis, S. M. et al. Evaluation of FluSight influenza forecasting in the 2021–22 and 2022–23 seasons with a new target laboratory-confirmed influenza hospitalizations. Nat. Commun. 15, 6289 (2024), doi:10.1038/s41467-024-50601-9.

31. Cramer, E. Y., et al. The United States COVID-19 Forecast Hub dataset. Sci. Data 9, 462 (2022), doi:10.1038/s41597-022-01517-w.

32. The Consortium of Infectious Disease Modeling Hubs. Hubverse Website. https://hubverse.io/.

33. Held, L., Meyer, S. & Bracher, J. Probabilistic forecasting in infectious disease epidemiology: the 13th Armitage lecture. Stat. Med. 36, 3443–3460 (2017), doi:10.1002/sim.7363.

34. Rumack, A., Tibshirani, R. J. & Rosenfeld, R. Recalibrating probabilistic forecasts of epidemics. *ArXiv211206305 Cs* (2021).

35. Raftery, A. E. Use and Communication of Probabilistic Forecasts. Stat. Anal. Data Min. 9, 397–410 (2016), doi:10.1002/sam.11302.

36. Krystalli, A. hubAdmin: Utilities for Administering Hubverse Hubs. R package version 1.6.0. https://github.com/hubverse-org/hubAdmin. (2025).

37. Krystalli, A., Ray, E., & Gruson, H. hubValidations: Testing framework for hubverse hub validations. R package version 0.11.0. https://github.com/hubverse-org/hubValidations. (2025).

38. Krystalli, A. hubData: Tools for accessing and working with hubverse data. R package version 1.3.0. https://github.com/hubverse-org/hubData. (2024).

39. Richardson, Neal et al. arrow: Integration to ‘Apache’ ‘Arrow’. R package version 19.0.1. https://arrow.apache.org/docs/r/ (2025).

40. Ray, E. L., et al. hubEnsembles: Ensemble Methods for Combining Hub Model Outputs, version 0.1.9. Comprehensive R Archive Network (2024).

41. Shandross, L. et al. Multi-Model Ensembles in Infectious Disease and Public Health: Methods, Interpretation, and Implementation in R. Stat. Med. 45, e70333 (2026), doi:10.1002/sim.70333.

42. Howerton, E. et al. Context-dependent representation of within- and between-model uncertainty: aggregating probabilistic predictions in infectious disease epidemiology. J. R. Soc. Interface 20, 20220659 (2023), doi:10.1098/rsif.2022.0659.

43. Contamin, L. hubVis: Plotting methods for hub models output. R package version 0.1.1. https://github.com/hubverse-org/hubVis. (2025).

44. Ray, E., Sweger, B., & Contamin, L. hubExamples: Example Hub Data. R package version 0.1.0. https://github.com/hubverse-org/hubExamples. (2025).

45. Reich, N., et al. hubEvals: Basic tools for scoring hubverse forecasts. R package version 0.0.0.9001. https://hubverse-org.github.io/hubEvals/. (2025).

46. CDC. CDC. MMWR Weeks Fact Sheet. https://ndc.services.cdc.gov/wp-content/uploads/MMWR_Week_overview.pdf. (2023).

47. Council of State and Territorial Epidemiologists. 2024 Epidemiology Capacity Assessment. https://www.cste.org/group/ECA (2024).

48. Wu, H. & Levinson, D. The ensemble approach to forecasting: A review and synthesis. Transp. Res. Part C Emerg. Technol. 132, 103357 (2021), doi:10.1016/j.trc.2021.103357.

49. Clemen, R. T. Combining forecasts: A review and annotated bibliography. Int. J. Forecast. 5, 559–583 (1989), doi:10.1016/0169-2070(89)90012-5.

50. Gneiting, T. & Raftery, A. E. Weather Forecasting with Ensemble Methods. Science 310, 248–249 (2005), doi:10.1126/science.1115255.

51. Hibon, M. & Evgeniou, T. To combine or not to combine: selecting among forecasts and their combinations. Int. J. Forecast. 21, 15–24 (2005), doi:10.1016/j.ijforecast.2004.05.002.

52. Polikar, R. Ensemble based systems in decision making. IEEE Circuits Syst. Mag. 6, 21–45 (2006), doi:10.1109/MCAS.2006.1688199.

53. Bosse, N. I. et al. Scoring epidemiological forecasts on transformed scales. PLOS Comput. Biol. 19, e1011393 (2023), doi:10.1371/journal.pcbi.1011393.

54. Loo, S. L. et al. Scenario Projections of COVID-19 Burden in the US, 2024-2025. JAMA Netw. Open 8, e2532469 (2025), doi:10.1001/jamanetworkopen.2025.32469.

55. Fox, S.J., Ryan, J, Salcedo, M, & Case, B. microhub: Forecasting tool for infectious disease modeling. https://github.com/sjfox/microhub. (2024).

56. The Apache Software Foundation. Apache Parquet. https://parquet.apache.org/ (2025).

57. R Core Team. R: A Language and Environment for Statistical Computing. https://www.R-project.org/. (2024).

58. Python Software Foundation. Python Language Reference, v. 3.13.5. https://www.python.org/. (2025).

