## Supplementary figures and images for "A Software Platform for Collaborative Infectious Disease Modeling"

### Supplementary figure 1

Flowchart of Hub Phases and the Tools Used in Each Phase

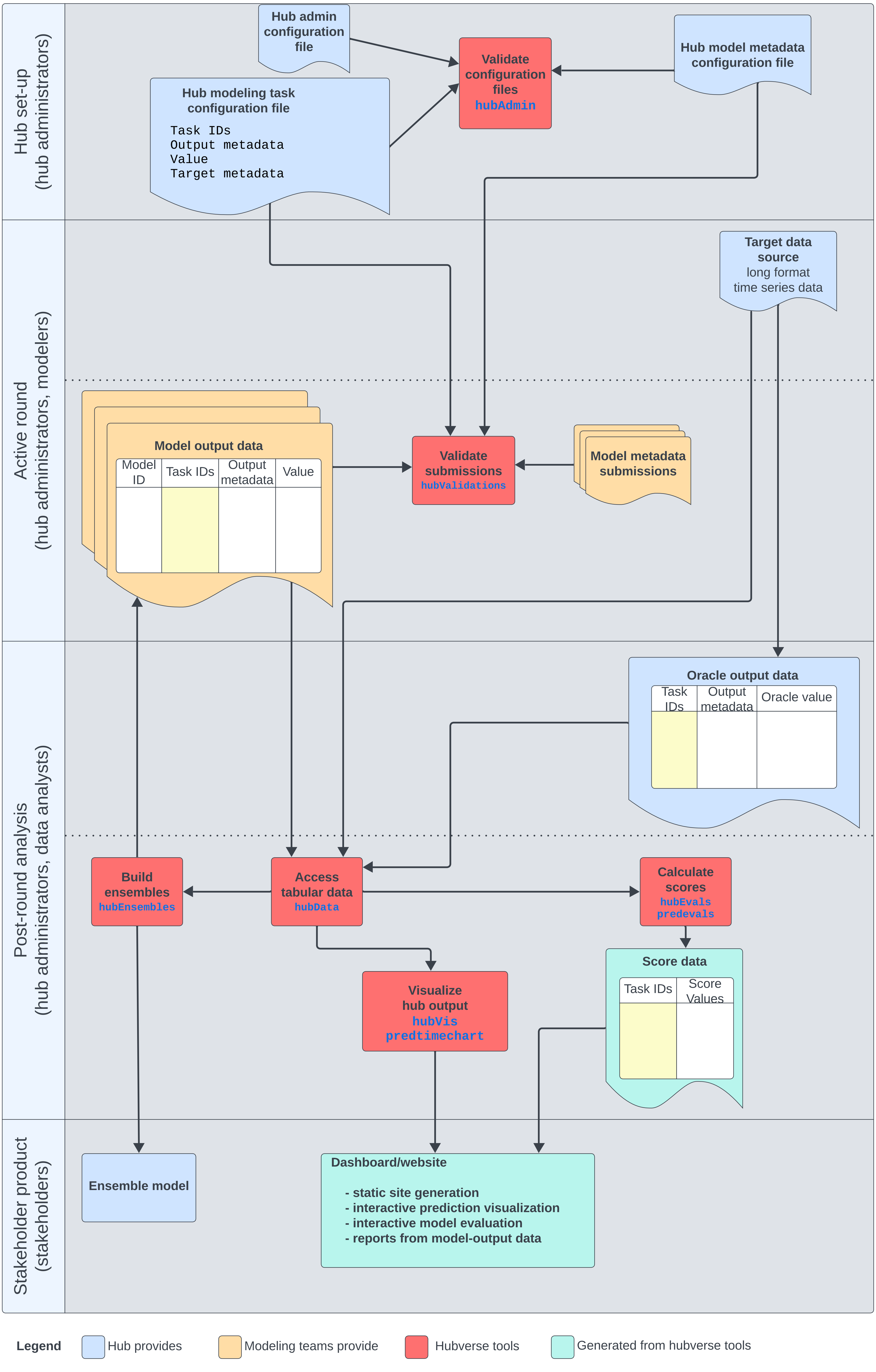
