## Supplementary data 1 for "A Software Platform for Collaborative Infectious Disease Modeling"

### FluSight Modeling Tasks Configuration File


```
{
    "schema_version": "https://raw.githubusercontent.com/Infectious-Disease-Modeling-
Hubs/schemas/main/v3.0.1/tasks-schema.json",
    "rounds": [
        {
            "round_id_from_variable": true,
            "round_id": "reference_date",
            "model_tasks": [
                {
                    "task_ids": {
                        "reference_date": {
                            "required": null,
                            "optional": [
                                "2023-10-07",
                                "2023-10-14",
                                "2023-10-21",
                                "2023-10-28",
                                "2023-11-04",
                                "2023-11-11",
                                "2023-11-18",
                                "2023-11-25",
                                "2023-12-02",
                                "2023-12-09",
                                "2023-12-16",
                                "2023-12-23",
                                "2023-12-30",
                                "2024-01-06",
                                "2024-01-13",
                                "2024-01-20",
                                "2024-01-27",
                                "2024-02-03",
                                "2024-02-10",
                                "2024-02-17",
                                "2024-02-24",
                                "2024-03-02",
                                "2024-03-09",
                                "2024-03-16",
                                "2024-03-23",
                                "2024-03-30",
                                "2024-04-06",
                                "2024-04-13",
                                "2024-04-20",
                                "2024-04-27",
                                "2024-05-04",
                                "2024-05-11"
                            ]
                        },
                        "target": {
                            "required": null,
                            "optional": [
                                "wk flu hosp rate change"
                            ]
                        },
                        "horizon": {
                            "required": null,
                            "optional": [
                                -1,
                                0,
                                1,
                                2,
                                3
                            ]
```

```
},
"location": {
    "required": null,
    "optional": [
        "US",
        "01",
        "02",
        "04",
        "05",
        "06",
        "08",
        "09",
        "10",
        "11",
        "12",
        "13",
        "15",
        "16",
        "17",
        "18",
        "19",
        "20",
        "21",
        "22",
        "23",
        "24",
        "25",
        "26",
        "27",
        "28",
        "29",
        "30",
        "31",
        "32",
        "33",
        "34",
        "35",
        "36",
        "37",
        "38",
        "39",
        "40",
        "41",
        "42",
        "44",
        "45",
        "46",
        "47",
        "48",
        "49",
        "50",
        "51",
        "53",
        "54",
        "55",
        "56",
        "72"
    ]
},
"target_end_date": {
    "required": null,
    "optional": [
        "2023-09-23",
        "2023-09-30",
        "2023-10-07",
        "2023-10-14",
        "2023-10-21",
```

"2023-10-28",
"2023-11-04",
"2023-11-11",
"2023-11-18",
"2023-11-25",
"2023-12-02",
"2023-12-09",
"2023-12-16",
"2023-12-23",
"2023-12-30",
"2024-01-06",
"2024-01-13",
"2024-01-20",
"2024-01-27",
"2024-02-03",
"2024-02-10",
"2024-02-17",
"2024-02-24",
"2024-03-02",
"2024-03-09",
"2024-03-16",
"2024-03-23",
"2024-03-30",
"2024-04-06",
"2024-04-13",
"2024-04-20",
"2024-04-27",
"2024-05-04",
"2024-05-11",
"2024-05-18",
"2024-05-25",
"2024-06-01"
]
}
},
"output_type": {
"pmf": {
"output_type_id": {
"required": [
"large_decrease",
"decrease",
"stable",
"increase",
"large_increase"
],
"optional": null
},
"value": {
"type": "double",
"minimum": 0,
"maximum": 1
}
}
},
"target_metadata": [
{
"target_id": "flu hosp rate change",
"target_name": "week ahead weekly influenza hospitalization rate change",
"target_units": "rate per 100,000 population",
"target_keys": {
"target": [
"wk flu hosp rate change"
]
},
"target_type": "ordinal",
"description": "This target represents the change in the rate of new

```
hospitalizations per week comparing the week ending on the reference_date to the week ending
[horizon] weeks after the reference_date, on target_end_date.",
                            "is_step_ahead": true,
                            "time_unit": "week"
                        }
                    ]
                },
                {
                    "task_ids": {
                        "reference_date": {
                            "required": null,
                            "optional": [
                                "2023-10-07",
                                "2023-10-14",
                                "2023-10-21",
                                "2023-10-28",
                                "2023-11-04",
                                "2023-11-11",
                                "2023-11-18",
                                "2023-11-25",
                                "2023-12-02",
                                "2023-12-09",
                                "2023-12-16",
                                "2023-12-23",
                                "2023-12-30",
                                "2024-01-06",
                                "2024-01-13",
                                "2024-01-20",
                                "2024-01-27",
                                "2024-02-03",
                                "2024-02-10",
                                "2024-02-17",
                                "2024-02-24",
                                "2024-03-02",
                                "2024-03-09",
                                "2024-03-16",
                                "2024-03-23",
                                "2024-03-30",
                                "2024-04-06",
                                "2024-04-13",
                                "2024-04-20",
                                "2024-04-27",
                                "2024-05-04",
                                "2024-05-11"
                            ]
                        },
                        "target": {
                            "required": null,
                            "optional": [
                                "wk inc flu hosp"
                            ]
                        },
                        "horizon": {
                            "required": null,
                            "optional": [
                                -1,
                                0,
                                1,
                                2,
                                3
                            ]
                        },
                        "location": {
                            "required": null,
                            "optional": [
                                "US",
                                "01",
```

```
            "02",
            "04",
            "05",
            "06",
            "08",
            "09",
            "10",
            "11",
            "12",
            "13",
            "15",
            "16",
            "17",
            "18",
            "19",
            "20",
            "21",
            "22",
            "23",
            "24",
            "25",
            "26",
            "27",
            "28",
            "29",
            "30",
            "31",
            "32",
            "33",
            "34",
            "35",
            "36",
            "37",
            "38",
            "39",
            "40",
            "41",
            "42",
            "44",
            "45",
            "46",
            "47",
            "48",
            "49",
            "50",
            "51",
            "53",
            "54",
            "55",
            "56",
            "72"
        ]
    },
    "target_end_date": {
        "required": null,
        "optional": [
            "2023-09-23",
            "2023-09-30",
            "2023-10-07",
            "2023-10-14",
            "2023-10-21",
            "2023-10-28",
            "2023-11-04",
            "2023-11-11",
            "2023-11-18",
            "2023-11-25",
            "2023-12-02",
```

```
            "2023-12-09",
            "2023-12-16",
            "2023-12-23",
            "2023-12-30",
            "2024-01-06",
            "2024-01-13",
            "2024-01-20",
            "2024-01-27",
            "2024-02-03",
            "2024-02-10",
            "2024-02-17",
            "2024-02-24",
            "2024-03-02",
            "2024-03-09",
            "2024-03-16",
            "2024-03-23",
            "2024-03-30",
            "2024-04-06",
            "2024-04-13",
            "2024-04-20",
            "2024-04-27",
            "2024-05-04",
            "2024-05-11",
            "2024-05-18",
            "2024-05-25",
            "2024-06-01"
        ]
    }
},
"output_type": {
    "quantile": {
        "output_type_id": {
            "required": [
                0.01,
                0.025,
                0.05,
                0.1,
                0.15,
                0.2,
                0.25,
                0.3,
                0.35,
                0.4,
                0.45,
                0.5,
                0.55,
                0.6,
                0.65,
                0.7,
                0.75,
                0.8,
                0.85,
                0.9,
                0.95,
                0.975,
                0.99
            ],
            "optional": null
        },
        "value": {
            "type": "double",
            "minimum": 0
        }
    },
    "sample": {
        "output_type_id_params": {
            "is_required": false,
```

```
                        "type": "integer",
                        "min_samples_per_task": 100,
                        "max_samples_per_task": 100,
                        "compound_taskid_set": [
                            "reference_date",
                            "location",
                            "target"
                        ]
                    },
                    "value": {
                        "type": "integer",
                        "minimum": 0
                    }
                }
            },
            "target_metadata": [
                {
                    "target_id": "wk inc flu hosp",
                    "target_name": "incident influenza hospitalizations",
                    "target_units": "count",
                    "target_keys": {
                        "target": [
                            "wk inc flu hosp"
                        ]
                    },
                    "target_type": "continuous",
                    "description": "This target represents the count of new hospitalizations
in the week ending on the date [horizon] weeks after the reference_date, on the target_end_date.",
                    "is_step_ahead": true,
                    "time_unit": "week"
                }
            ]
        },
        {
            "task_ids": {
                "reference_date": {
                    "required": null,
                    "optional": [
                        "2023-10-07",
                        "2023-10-14",
                        "2023-10-21",
                        "2023-10-28",
                        "2023-11-04",
                        "2023-11-11",
                        "2023-11-18",
                        "2023-11-25",
                        "2023-12-02",
                        "2023-12-09",
                        "2023-12-16",
                        "2023-12-23",
                        "2023-12-30",
                        "2024-01-06",
                        "2024-01-13",
                        "2024-01-20",
                        "2024-01-27",
                        "2024-02-03",
                        "2024-02-10",
                        "2024-02-17",
                        "2024-02-24",
                        "2024-03-02",
                        "2024-03-09",
                        "2024-03-16",
                        "2024-03-23",
                        "2024-03-30",
                        "2024-04-06",
                        "2024-04-13",
                        "2024-04-20",
```

```
        "2024-04-27",
        "2024-05-04",
        "2024-05-11"
    ]
},
"target": {
    "required": null,
    "optional": [
        "peak inc flu hosp"
    ]
},
"horizon": {
    "required": null,
    "optional": null
},
"location": {
    "required": null,
    "optional": [
        "US",
        "01",
        "02",
        "04",
        "05",
        "06",
        "08",
        "09",
        "10",
        "11",
        "12",
        "13",
        "15",
        "16",
        "17",
        "18",
        "19",
        "20",
        "21",
        "22",
        "23",
        "24",
        "25",
        "26",
        "27",
        "28",
        "29",
        "30",
        "31",
        "32",
        "33",
        "34",
        "35",
        "36",
        "37",
        "38",
        "39",
        "40",
        "41",
        "42",
        "44",
        "45",
        "46",
        "47",
        "48",
        "49",
        "50",
        "51",
        "53",
```

```
                        "54",
                        "55",
                        "56",
                        "72"
                    ]
                },
                "target_end_date": {
                    "required": null,
                    "optional": null
                }
            },
            "output_type": {
                "quantile": {
                    "output_type_id": {
                        "required": [
                            0.01,
                            0.025,
                            0.05,
                            0.1,
                            0.15,
                            0.2,
                            0.25,
                            0.3,
                            0.35,
                            0.4,
                            0.45,
                            0.5,
                            0.55,
                            0.6,
                            0.65,
                            0.7,
                            0.75,
                            0.8,
                            0.85,
                            0.9,
                            0.95,
                            0.975,
                            0.99
                        ],
                        "optional": null
                    },
                    "value": {
                        "type": "double",
                        "minimum": 0
                    }
                }
            },
            "target_metadata": [
                {
                    "target_id": "peak inc flu hosp",
                    "target_name": "incident influenza hospitalizations in the season week
with the highest hospitalizations",
                    "target_units": "count",
                    "target_keys": {
                        "target": [
                            "peak inc flu hosp"
                        ]
                    },
                    "target_type": "continuous",
                    "description": "This target represents the count of new hospitalizations
 in the week of the season with the highest reported hospitalizations.",
                    "is_step_ahead": false
                }
            ]
        },
        {
            "task_ids": {
```

```
"reference_date": {
    "required": null,
    "optional": [
        "2023-10-07",
        "2023-10-14",
        "2023-10-21",
        "2023-10-28",
        "2023-11-04",
        "2023-11-11",
        "2023-11-18",
        "2023-11-25",
        "2023-12-02",
        "2023-12-09",
        "2023-12-16",
        "2023-12-23",
        "2023-12-30",
        "2024-01-06",
        "2024-01-13",
        "2024-01-20",
        "2024-01-27",
        "2024-02-03",
        "2024-02-10",
        "2024-02-17",
        "2024-02-24",
        "2024-03-02",
        "2024-03-09",
        "2024-03-16",
        "2024-03-23",
        "2024-03-30",
        "2024-04-06",
        "2024-04-13",
        "2024-04-20",
        "2024-04-27",
        "2024-05-04",
        "2024-05-11"
    ]
},
"target": {
    "required": null,
    "optional": [
        "peak week inc flu hosp"
    ]
},
"horizon": {
    "required": null,
    "optional": null
},
"location": {
    "required": null,
    "optional": [
        "US",
        "01",
        "02",
        "04",
        "05",
        "06",
        "08",
        "09",
        "10",
        "11",
        "12",
        "13",
        "15",
        "16",
        "17",
        "18",
        "19",
```

```
                "20",
                "21",
                "22",
                "23",
                "24",
                "25",
                "26",
                "27",
                "28",
                "29",
                "30",
                "31",
                "32",
                "33",
                "34",
                "35",
                "36",
                "37",
                "38",
                "39",
                "40",
                "41",
                "42",
                "44",
                "45",
                "46",
                "47",
                "48",
                "49",
                "50",
                "51",
                "53",
                "54",
                "55",
                "56",
                "72"
            ]
        },
        "target_end_date": {
            "required": null,
            "optional": null
        }
    },
    "output_type": {
        "pmf": {
            "output_type_id": {
                "required": [
                    "2024-11-09",
                    "2024-11-16",
                    "2024-11-23",
                    "2024-11-30",
                    "2024-12-07",
                    "2024-12-14",
                    "2024-12-21",
                    "2024-12-28",
                    "2025-01-04",
                    "2025-01-11",
                    "2025-01-18",
                    "2025-01-25",
                    "2025-02-01",
                    "2025-02-08",
                    "2025-02-15",
                    "2025-02-22",
                    "2025-03-01",
                    "2025-03-08",
                    "2025-03-15",
                    "2025-03-22",
```

```
                                        "2025-03-29",
                                        "2025-04-05",
                                        "2025-04-12",
                                        "2025-04-19",
                                        "2025-04-26",
                                        "2025-05-03",
                                        "2025-05-10",
                                        "2025-05-17",
                                        "2025-05-24",
                                        "2025-05-31"
                                    ],
                                    "optional": null
                                },
                                "value": {
                                    "type": "double",
                                    "minimum": 0,
                                    "maximum": 1
                                }
                            }
                        },
                        "target_metadata": [
                            {
                                "target_id": "peak week inc flu hosp",
                                "target_name": "season week with the highest hospitalizations",
                                "target_units": "count",
                                "target_keys": {
                                    "target": [
                                        "peak week inc flu hosp"
                                    ]
                                },
                                "target_type": "date",
                                "description": "This target represents the week of the season with the
highest reported hospitalizations. This is the Saturday ending the epidemic week with the highest
reported hospitalization count.",
                                "is_step_ahead": false
                            }
                        ]
                    }
                ],
                "submissions_due": {
                    "relative_to": "reference_date",
                    "start": -6,
                    "end": -3
                }
            }
        ]
}
```
