## Supplementary note 1 for "A Software Platform for Collaborative Infectious Disease Modeling"

#### Accessing Data from the US Flusight Hubverse Project

2025-07-02

### Introduction

This vignette serves as supplementary material for the manuscript "Coordinating collaborative infectious disease modeling projects with the hubverse". The following text appears in the manuscript to introduce the motivating example presented here:

> The FluSight challenge, run annually by U.S. CDC since 2013 (with a one-year break during the COVID-19 pandemic), has served as a centrally coordinated collaborative effort to monitor and predict short-term influenza activity in the U.S. at the national, regional, and state level (https://github.com/cdcepi/FluSight-forecast-hub/). These seasonal collaborative forecasting challenges have garnered participation from dozens of academic, industry, and governmental research teams. Typically, forecasts have been submitted once a week from October through May (the typical influenza season in the U.S.). Forecasts consisted of quantitative predictions of observed values from public health surveillance systems that are consistently reported across the country. The exact data source and prediction targets have changed over the duration of the project. Starting in the 2023-2024 season, FluSight has used the hubverse ecosystem to configure and maintain submissions of short-term forecasts (https://github.com/cdcepi/FluSight-forecast-hub/releases/tag/v1.0.0).

We refer readers to the main manuscript or the hubverse website to learn more about the key concepts and definitions of terms that underlie this demonstration.

### Required software

The following demonstration shows how to access and analyze data from a real hub using and several of the hubverse R packages[The Consortium of Infectious Disease Modeling Hubs, 2025].

The hubverse packages `hubData`, `hubVis`, `hubEnsembles` and `hubEvals` are best installed through the R-universe package repository to ensure you are working with the latest released versions. Note that these packages have some dependencies that are needed for the operations in this vignette, so specifying `dependencies = TRUE` is recommended. The packages can be installed from the R console using the following command:

```
install.packages(
  pkgs = c("hubData", "hubVis", "hubEnsembles", "hubEvals"),
  repos = c(
    "https://hubverse-org.r-universe.dev",
    "https://cloud.r-project.org"
```

```r
  ),
  dependencies = TRUE
)
```

Supporting packages `ggplot2` and `dplyr` can be installed from CRAN.

```r
install.packages(
  pkgs = c("dplyr", "ggplot2")
)
```

Once the required packages are installed, load the supporting packages into your R session. (Session information on all the packages used including package versions are included at the end of this vignette.)

```r
# Data munging
library(dplyr)
# Plotting
library(ggplot2)
```

#### Load forecast data

CDC's FluSight Forecast Hub exists as a GitHub repository[US CDC, 2025]. Using hubverse tools, the data for this hub has been validated and mirrored to the cloud, so a copy of the model output for this hub lives at a publicly-accessible AWS S3 bucket. This means that instead of needing to download a local copy of the repository, the data for this project can be directly accessed from the cloud using `hubData` tools. The data is organised in a format we can open as an arrow dataset through R package `arrow` directly .

We start by initializing a connection to the hub in the cloud. Printing displays metadata about the resulting connection.

```r
hub_path <- hubData::s3_bucket("cdcepi-flusight-forecast-hub/")
hub_con <- hubData::connect_hub(hub_path, skip_checks = TRUE)
hub_con
#>
#> -- <hub_connection/FileSystemDataset> --
#>
#> * hub_name: "US CDC FluSight"
#> * hub_path: 'cdcepi-flusight-forecast-hub/'
#> * file_format: "parquet(2454/2454)"
#> * checks: FALSE
#> * file_system: "S3FileSystem"
#> * model_output_dir: "model-output/"
#> * config_admin: 'hub-config/admin.json'
#> * config_tasks: 'hub-config/tasks.json'
#>
#> -- Connection schema
#> hub_connection with 2454 Parquet files
#> 9 columns
#> reference_date: date32[day]
#> target: string
#> horizon: int32
#> location: string
#> target_end_date: date32[day]
#> output_type: string
#> output_type_id: string
#> value: double
```

```
#> model_id: string
```

The hub connection object printed above shows information about the hub, including the number and format of files loaded, the schema of the data, and column names and data types of model-output data. Note that the `skip_checks = TRUE` argument is used to skip the validation checks that are normally performed when connecting to a hub. In practice, since these data are all validated upon submission to the hub and since the checks impact performance, we omit the checks in this example.

We then collect a subset of model outputs that we will interact with further in this demonstration. Specifically, we will extract zero through three week-ahead quantile predictions of weekly incident flu hospitalizations made for the state of Texas on three dates in late 2024 and early 2025. For simplicity, we will restrict our query to predictions made for five arbitrarily chosen models, although predictions for over 30 models are available on each of these dates. Although the data are stored in the hub across multiple files, organised by `model_id` and `round_id`, `hubData::connect_hub()` wraps functionality from `arrow` to open a connection to the data as an Arrow Dataset. This allows us to act on all of the data collectively, as if it were a single `data.frame` object. Arrow datasets, including those stored on S3, behave as lazy data frames—they do not load data into memory until explicitly queried or collected. The `arrow` package also provides a dplyr interface to Arrow Datasets which allows us to query and manipulate the data.

```
models <- c(
  "FluSight-baseline",
  "CMU-TimeSeries",
  "UMass-flusion",
  "cfa-flumech",
  "UGA_flucast-INFLAenza"
)

reference_dates <- c("2024-12-14", "2025-01-11", "2025-02-08")

flusight_data <- hub_con |>
  filter(
    target == "wk inc flu hosp",
    reference_date %in% reference_dates,
    model_id %in% models,
    output_type == "quantile",
    location == "48", ## FIPS code for Texas
    horizon > -1
  ) |>
  hubData::collect_hub()
```

The `collect_hub` `hubData` function wraps the `dplyr::collect()` function but also coerces the output to a `model_out_tbl` class object where possible.

Table 1: Five rows of the `flusight_data` object collected above. The “target”, “location”, and “target_end_date” columns have been excluded for readability. The “output_type_id” and “value” columns show the different predicted quantiles for the number of hospital admissions due to influenza made by the CMU-TimeSeries model for location 48 (Texas). The predictions were made during the week of December 14, 2024 (reference date) and are making a prediction for the horizon of 1 week ahead, for the week ending December 21.

| model_id | reference_date | horizon | output_type | output_type_id | value |
|---|---|---|---|---|---|
| CMU-TimeSeries | 2025-01-11 | 1 | quantile | 0.01 | 304.41 |
| CMU-TimeSeries | 2025-01-11 | 1 | quantile | 0.025 | 425.46 |
| CMU-TimeSeries | 2025-01-11 | 1 | quantile | 0.05 | 565.22 |
| CMU-TimeSeries | 2025-01-11 | 1 | quantile | 0.1 | 800.02 |
| CMU-TimeSeries | 2025-01-11 | 1 | quantile | 0.15 | 1013.59 |

Let’s also create a vector of target_end_dates that we will use later to filter the oracle-output target data. The `target_end_date` column in the model output data indicates the date for which the prediction is made, and is used to align model output data with target data. In model output data it is defined as `reference_date` + `horizon * 7L` (in weeks).

```
target_end_dates <- unique(flusight_data$target_end_date)
```

#### Loading target data

To be able to visualize forecasts in relation to observed data, we need to load the target (observed) data for the locations and time period we are interested in.

Hubs generally contain two types of target data: time-series data and oracle output data. Time-series data is stored in a long format, with one row per observation, a single observed value per observation unit and is the format used for visualization of observed time-series target data. Oracle output data on the other hand is often derived from time-series data but more closely matches the model output data format and is used for the evaluation of model outputs. The FluSight hub stores target data in both formats, but we will first load the time-series format to visualize the performance of hub models against observed data. For more information on target data formats, see the hubverse documentation.

Accessing target data follows a similar pattern to accessing model output data. We start by connecting to the target data type of interest in the hub. To access the time-series format we use `hubData::connect_target_timeseries()` function.

```
ts_data_con <- hubData::connect_target_timeseries(hub_path)
ts_data_con
#> target_timeseries with 1 csv file
#> 7 columns
#> as_of: date32[day]
#> target: string
#> target_end_date: date32[day]
#> location: string
#> location_name: string
#> observation: double
#> weekly_rate: double
```

Next, we filter the target data to the location and time period of interest. We want to plot a time-series of observed data for the entire season. So, we are interested in the state of Texas (FIPS code 48) and the 2024-2025 respiratory virus season, which runs from September 23, 2024 to May 1, 2024. We also filter to the most recent `as_of` date to ensure we are using the latest available data. The `as_of` date is a metadata field that indicates when the target data was last updated.

To enable filtering by the maximum (latest) value of `as_of`, we use the `arrow::to_duckdb()` function to convert the connection to a memory virtual DuckDB table. This allows us to run `dplyr`-style queries that are supported by DuckDB but not by arrow, including filtering for the maximum value of a column. We then filter the data to the desired location, time period, and maximum `as_of` date, and select the relevant columns. Finally, we collect the data into a local data frame and rename the `target_end_date` column to `date` which is what the `hubVis` package functions expect.

```
target_data <- ts_data_con |>
  arrow::to_duckdb() |>
  filter(
    location == "48",
    target_end_date >= "2024-09-23",
    target_end_date <= "2025-05-01",
    as_of == max(as_of)
  ) |>
  select(location, location_name, target, target_end_date, observation) |>
  arrange(target_end_date) |>
  collect()
```

The resulting target data contains the actual number of hospital admissions due to influenza in Texas for the 2024-2025 respiratory virus season, which is the same target that hub models the models are predicting.

Table 2: Five rows of the time-series `target_data` object read in above. The observation column shows the number of hospital admissions due to influenza reported in Texas (location 48) on the week ending on each date.

| location | location_name | target | target_end_date | observation |
|---|---|---|---|---|
| 48 | Texas | wk inc flu hosp | 2024-09-28 | 1 |
| 48 | Texas | wk inc flu hosp | 2024-10-05 | 1 |
| 48 | Texas | wk inc flu hosp | 2024-10-12 | 79 |
| 48 | Texas | wk inc flu hosp | 2024-10-19 | 46 |
| 48 | Texas | wk inc flu hosp | 2024-10-26 | 63 |

#### Visualize forecasts

A common analysis task when working with forecast data is visualization. Plotting forecasts against the eventual observations provides a qualitative evaluation tool (visual inspection), and can often assist in diagnosing problems with models. We can use the hubVis package to create a single graphic that overlays the forecasts from the five models and three forecast dates with the observed data.

First we set the default theme for plots to be a clean black and white theme.

```
theme_set(theme_bw())
```

Next we use function `plot_step_ahead_model_output()` to visualize quantile, mean and median model output data in relation to target data. This function takes the model output data and target data as input, and plots the predictions from each model along with the observed data. The `x_col_name` argument specifies the column to use for the x-axis, which is `target_end_date` in this case. The `use_median_as_point` argument specifies whether to use the median prediction as the point estimate for each model, and `group` specifies how to group the data for plotting. The `fill_transparency` argument controls the transparency of the shaded prediction intervals, and `intervals` specifies the width of the prediction intervals to plot.

```
flusight_data |>
  hubVis::plot_step_ahead_model_output(
    target_data,
    x_col_name = "target_end_date",
    x_target_col_name = "target_end_date",
    use_median_as_point = TRUE,
    group = "reference_date",
    interactive = FALSE,
    fill_transparency = .1,
    intervals = 0.8
  ) +
  theme(legend.position = "bottom") +
  ylab("hospital admissions due to influenza")
```

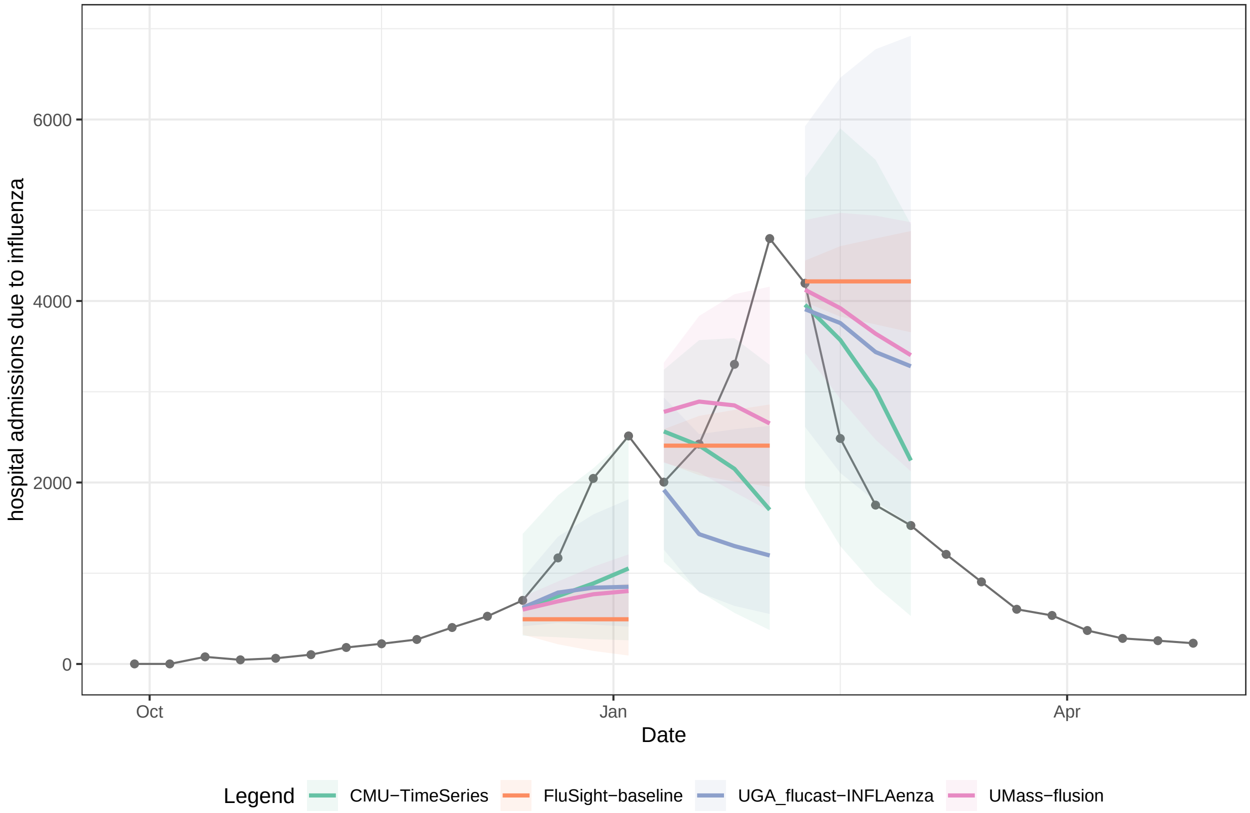


Figure 1: Forecasts of hospital admissions in Texas due to influenza in the 2024-2025 respiratory virus season. Predictions from five models are shown along with observed data (line with dots for specific observations). Each model is shown in a different color, with the 80% prediction intervals shown as a shaded region.

#### Build ensemble and visualize

Ensemble forecasting is the practice of combining predictions from multiple models to improve forecast accuracy. One of the advantages of using a coordinated modeling hub is that it can make it easier to combine predictions from multiple models, as the hub provides a standardized format for model output data[Reich et al., 2022]. The `hubEnsembles` package provides tools to build ensembles from hubverse-style model output data.

Using the model output data collected above, we can pass these data to the `hubEnsembles::simple_ensemble()` function to build an ensemble that takes the mean of predicted values from each model at each quantile level. This new forecast output is then added to the existing forecasts. The `hubEnsembles` package supports a range of aggregation approaches including a linear opinion pool (probability density averaging) or using the mean or median of quantiles[Shandross et al., 2025]. Model weights can be specified for the various approaches, allowing for some models to influence the output more strongly.

```
ensemble_forecast <- hubEnsembles::simple_ensemble(flusight_data)

all_forecasts <- flusight_data |>
  bind_rows(ensemble_forecast)
```

We can now visualize the ensemble forecast as well as individual model performance all together.

```
all_forecasts |>
  hubVis::plot_step_ahead_model_output(target_data,
    x_col_name = "target_end_date",
    x_target_col_name = "target_end_date",
    use_median_as_point = TRUE,
    group = "reference_date",
    interactive = FALSE,
    fill_transparency = .2,
    intervals = 0.8,
    one_color = "darkgray",
    ens_name = "hub-ensemble",
    ens_color = "blue",
    pal_color = NULL
  ) +
  theme(legend.position = "none") +
  ylab("hospital admissions due to influenza")
```

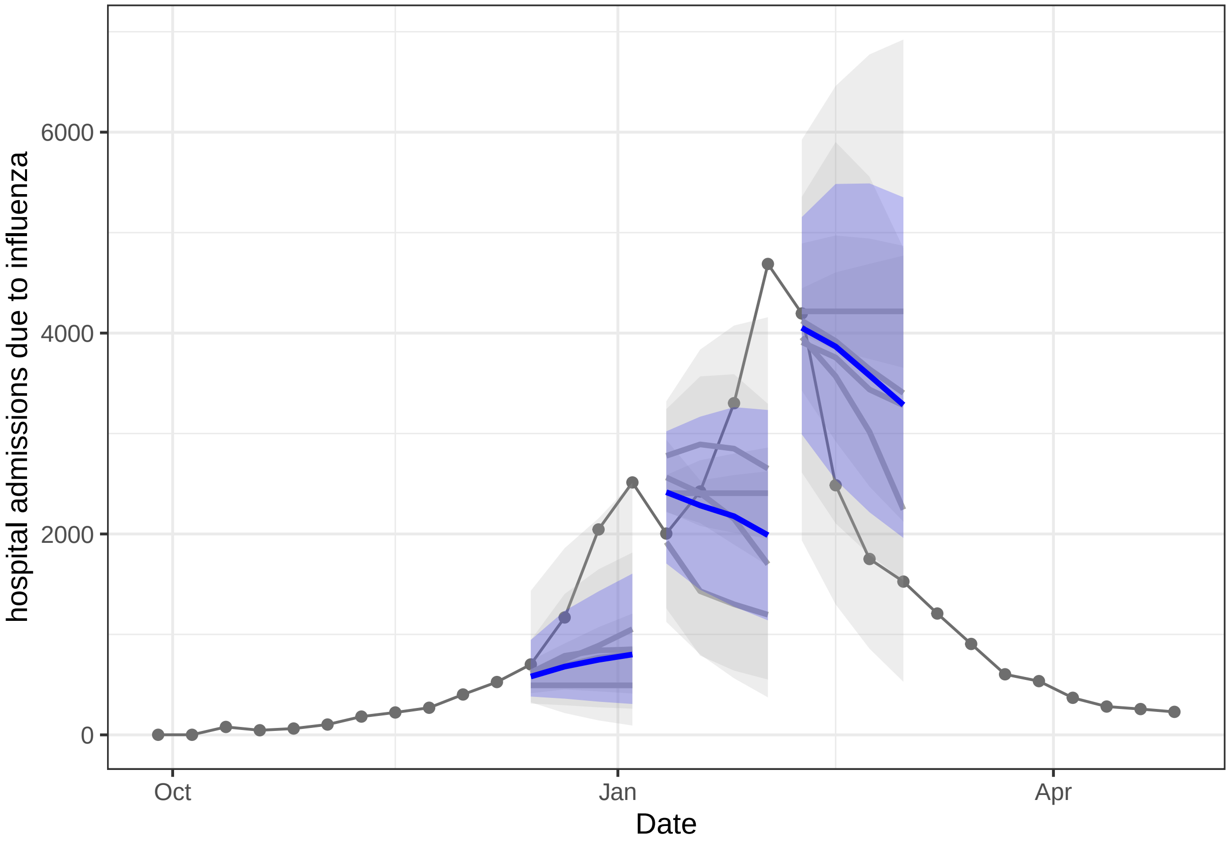
hospital admissions due to influenza
6000
4000
2000
0
Oct
Jan
Apr
Date

#### Evaluate forecasts

In many settings, point and probabilistic forecasts can be compared with eventual observations to provide a quantitative evaluation of model performance. These evaluations can provide insights into the comparative accuracy of different forecasting approaches, and can help identify areas for improvement in the models.

The package `hubEvals` can be used to evaluate probabilistic predictions against observed data. The `hubEvals` package creates an interface for hubverse-style model output and oracle output target data to the `scoringutils` package, which is an engine for computing a number of common scores for probabilistic predictions[Bosse et al., 2024].

As mentioned earlier, to evaluate model outputs we use oracle output data, a form of target data which matches and can be merged with hubverse-style model output data but which contains a point prediction representing the eventual observation, as if observed by an oracle model.

The FluSight hub also stores oracle output data in the cloud, and we can access it using the `hubData::connect_target_oracle_output()` function. This function connects to the oracle output data in the hub and returns a connection object that can again be used to query the data.

```
oo_data_con <- hubData::connect_target_oracle_output(hub_path)
oo_data_con
#> target_oracle_output with 1 csv file
#> 8 columns
#> as_of: date32[day]
#> target: string
#> target_end_date: date32[day]
#> location: string
#> horizon: int32
#> output_type: string
#> output_type_id: string
#> oracle_value: double
```

Once we have a connection to the data, we can query it for the data of interest. In this case, we want to filter the oracle output data to the same location and time period as the model output data, and for the same target.

```
oracle_output <- oo_data_con |>
  filter(
    target == "wk inc flu hosp",
    target_end_date %in% target_end_dates,
    output_type == "quantile",
    location == "48" ## FIPS code for Texas
  ) |>
  select(target, target_end_date, location, output_type, output_type_id, oracle_value) |>
  distinct() |>
  arrange(target_end_date) |>
  collect()
```

Table 3: Five rows of the `oracle_output` object from above. The `oracle_value` column shows the observed number of hospital admissions due to influenza reported in Texas (location 48) on the week ending on each `target_end_date`.

| target | target_end_date | location | output_type | oracle_value |
|---|---|---|---|---|
| wk inc flu hosp | 2024-12-14 | 48 | quantile | 701 |
| wk inc flu hosp | 2024-12-21 | 48 | quantile | 1169 |
| wk inc flu hosp | 2024-12-28 | 48 | quantile | 2045 |

| target | target_end_date | location | output_type | oracle_value |
|---|---|---|---|---|
| wk inc flu hosp | 2025-01-04 | 48 | quantile | 2513 |
| wk inc flu hosp | 2025-01-11 | 48 | quantile | 2004 |

Now that we have our oracle (observed) data in a format that can be combined with model output data, we can move on to evaluating it. In this example, we compute and show results for the average weighted interval score (WIS) with its associated decomposition into overprediction, underprediction, and dispersion penalties, the 50% and 90% prediction interval coverage rates, and the absolute error computed on the median prediction[Bracher et al., 2021]. To do this, we use the `hubEvals::score_model_out()` function which scores model outputs against observed data. Further information is available in the help page of the `hubEvals` package.

```
scores <- hubEvals::score_model_out(all_forecasts,
  oracle_output,
  baseline = "FluSight-baseline"
) |>
  arrange(wis)

scoringutils::plot_wis(scores, x = "model_id")
```

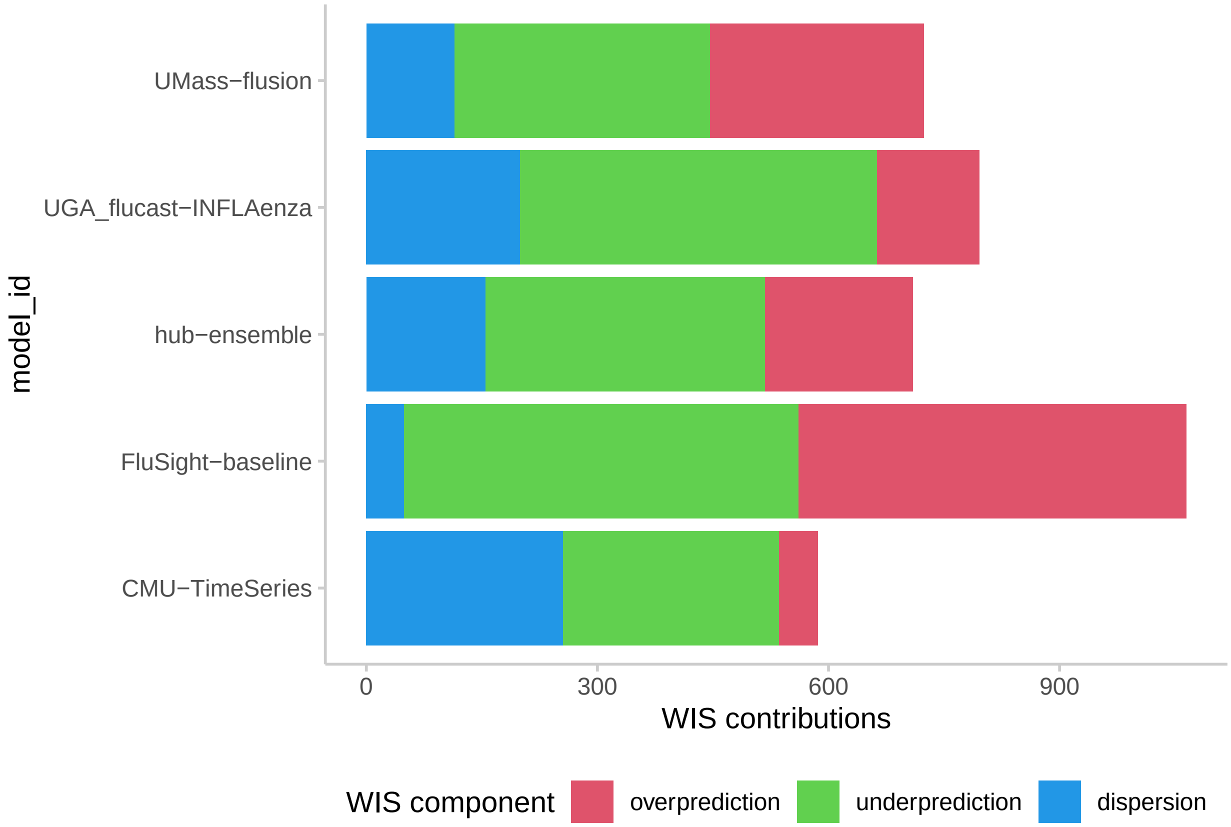


Figure 2: Average weighted interval score (WIS) computed by model (y-axis). The total length of each bar is the average WIS across all predictions made. WIS can be decomposed into penalties for overprediction, underprediction and dispersion, the contributions of which are shown by the segments of the bar. The UMass-flusion model has the lowest (best) average WIS, and it received a greater penalty for underprediction relative to the penalty for overprediction.

Table 4: A print-out of the `scores` object computed above, containing the summary metrics for the five component models and the ensemble model. Columns including other metrics, including bias, underprediction, overprediction and dispersion were omitted for space. Smaller values of WIS indicate more accurate performance. Interval coverage rates should be close to the nominal level of the interval (e.g., 0.5 for the 50% interval, 0.9 for the 90% interval).

| model_id | wis | interval_coverage_50 | interval_coverage_90 | ae_median |
|---|---:|---:|---:|---:|
| CMU-TimeSeries | 586.07 | 0.67 | 0.92 | 928.86 |
| hub-ensemble | 709.92 | 0.25 | 0.58 | 1092.22 |
| UMass-flusion | 723.83 | 0.17 | 0.50 | 1047.54 |
| UGA_flucast-INFLAenza | 795.60 | 0.25 | 0.67 | 1241.58 |
| FluSight-baseline | 1064.63 | 0.17 | 0.25 | 1246.67 |

#### Session information

```
sessionInfo()
#> R version 4.4.3 (2025-02-28)
#> Platform: aarch64-apple-darwin20
#> Running under: macOS Sequoia 15.5
#>
#> Matrix products: default
#> BLAS:   /Library/Frameworks/R.framework/Versions/4.4-arm64/Resources/lib/libRblas.0.dylib
#> LAPACK: /Library/Frameworks/R.framework/Versions/4.4-arm64/Resources/lib/libRlapack.dylib;  LAPACK v
#>
#> locale:
#> [1] en_US.UTF-8/en_US.UTF-8/en_US.UTF-8/C/en_US.UTF-8/en_US.UTF-8
#>
#> time zone: America/New_York
#> tzcode source: internal
#>
#> attached base packages:
#> [1] stats     graphics  grDevices utils     datasets  methods   base
#>
#> other attached packages:
#> [1] ggplot2_3.5.2 dplyr_1.1.4
#>
#> loaded via a namespace (and not attached):
#>  [1] gtable_0.3.6           xfun_0.52              htmlwidgets_1.6.4
#>  [4] gh_1.5.0               tzdb_0.5.0             vctrs_0.6.5
#>  [7] tools_4.4.3            generics_0.1.4         curl_6.3.0
#> [10] tibble_3.3.0           blob_1.2.4             pkgconfig_2.0.3
#> [13] data.table_1.17.4      checkmate_2.3.2        dbplyr_2.5.0
#> [16] RColorBrewer_1.1-3     assertthat_0.2.1       lifecycle_1.0.4
#> [19] compiler_4.4.3         farver_2.1.2           stringr_1.5.1
#> [22] tinytex_0.57           htmltools_0.5.8.1      yaml_2.3.10
#> [25] lazyeval_0.2.2         plotly_4.10.4          pillar_1.10.2
#> [28] tidyr_1.3.1            MASS_7.3-64            hubVis_0.1.2.9000
#> [31] hubEvals_0.0.0.9001    cachem_1.1.0           Metrics_0.1.4
#> [34] tidyselect_1.2.1       digest_0.6.37          hubData_1.4.0
#> [37] stringi_1.8.7          zoltr_1.0.2            duckdb_1.3.1
#> [40] purrr_1.0.4            labeling_0.4.3         arrow_20.0.0.2
#> [43] fastmap_1.2.0          grid_4.4.3             cli_3.6.5
#> [46] magrittr_2.0.3         hubUtils_0.5.0.9000    dichromat_2.0-0.1
#> [49] readr_2.1.5            withr_3.0.2            scales_1.4.0
#> [52] backports_1.5.0        bit64_4.6.0-1          rmarkdown_2.29
#> [55] httr_1.4.7             matrixStats_1.5.0      bit_4.6.0
#> [58] scoringutils_2.1.0.9000 hms_1.1.3             memoise_2.0.1
#> [61] evaluate_1.0.3         knitr_1.50             viridisLite_0.4.2
#> [64] scoringRules_1.1.3     rlang_1.1.6            Rcpp_1.0.14
#> [67] glue_1.8.0             DBI_1.2.3              hubEnsembles_1.0.0
#> [70] rstudioapi_0.17.1      jsonlite_2.0.0         R6_2.6.1
#> [73] fs_1.6.6
```

### References


Nikos I. Bosse, Hugo Gruson, Anne Cori, Edwin van Leeuwen, Sebastian Funk, and Sam Abbott. Evaluating forecasts with scoringutils in r. *arXiv*, 2024. doi: 10.48550/ARXIV.2205.07090. URL https://arxiv.org/abs/2205.07090.

Johannes Bracher, Evan L. Ray, Tilmann Gneiting, and Nicholas G. Reich. Evaluating epidemic forecasts in an interval format. *PLoS Computational Biology*, 17(2):e1008618, February 2021. doi: 10.1371/journal.pcbi.1008618. URL https://www.ncbi.nlm.nih.gov/pmc/articles/PMC7880475/.

Nicholas G. Reich, Justin Lessler, Sebastian Funk, Cecile Viboud, Alessandro Vespignani, Ryan J. Tibshirani, Katriona Shea, Melanie Schienle, Michael C. Runge, Roni Rosenfeld, Evan L. Ray, Rene Niehus, Helen C. Johnson, Michael A. Johansson, Harry Hochheiser, Lauren Gardner, Johannes Bracher, Rebecca K. Borchering, and Matthew Biggerstaff. Collaborative Hubs: Making the Most of Predictive Epidemic Modeling. *American Journal of Public Health*, 112(6):839–842, June 2022. ISSN 1541-0048. doi: 10.2105/AJPH.2022.306831.

Li Shandross, Emily Howerton, Lucie Contamin, Harry Hochheiser, Anna Krystalli, Consortium of Infectious Disease Modeling Hubs, Nicholas G. Reich, and Evan L. Ray. Multi-model ensembles in infectious disease and public health: Methods, interpretation, and implementation in R. May 2025. doi: 10.1101/2024.06.24.24309416. URL https://www.medrxiv.org/content/10.1101/2024.06.24.24309416v2.

The Consortium of Infectious Disease Modeling Hubs. *The hubverse: open tools for collaborative modeling.*, 2025. URL https://hubverse.io. Accessed: 2025-06-10.

US CDC. *cdcepi/FluSight-forecast-hub*, June 2025. URL https://github.com/cdcepi/FluSight-forecast-hub. Accessed: 2025-06-10.
